# ATRX loss in sarcomas is associated with dysregulated gene and transposable element expression, loss of DNA methylation, and worse survival

**DOI:** 10.1101/2025.11.13.25340146

**Authors:** Kathryn Demanelis, Julia Leix, Melissa A. Burgess, Benjamin A. Nacev

## Abstract

**Background:** *ATRX* is a tumor suppressor and key regulator of transcriptionally repressive epigenetic states. We investigate the impact of ATRX loss on gene expression, DNA methylation, and transposable element (TE) expression in common soft tissue sarcoma tumors, a family of rare cancers.

**Methods:** Using data from The Cancer Genome Atlas (TCGA), we analyzed 234 tumors from patients with dedifferentiated liposarcoma (DDLPS), undifferentiated pleomorphic sarcoma (UPS), soft tissue leiomyosarcoma (LMS), uterine LMS (uLMS), and myxofibrosarcoma (MFS). Corresponding clinical outcome and patient data, DNA sequencing, RNA sequencing, and 450K methylation data were integrated to assess ATRX-dependent features.

**Results:** ATRX loss was associated with significantly altered gene expression with the greatest impact in LMS/uLMS with 31 (33.0%) genes downregulated and 63 (66.0%) genes upregulated (FDR < 0.05), consistent with a role for ATRX in transcriptional silencing. Methylation profiling identified 269 differentially methylated CpGs (FDR<0.05), with a marked hypomethylation effect. ATRX loss was associated with loss of TE silencing with 97% (n=93) of differentially expressed TEs upregulated, indicating that ATRX loss contributes to a marked de-repression of TEs. The median overall survival in LMS patients was 81.0 vs 34.9 months for tumors with ATRX retention and loss (p_log-rank_=0.0023).

**Conclusions:** ATRX loss in sarcomas leads to DNA hypomethylation, increased expression of TEs, as well as transcriptional dysregulation affecting key oncogenic pathways. ATRX status may serve as a potential biomarker for prognosis and therapeutic stratification. Future clinical trials investigating epigenetic therapies could offer novel treatment strategies for ATRX-deficient sarcomas.

## Introduction

Sarcomas are a heterogeneous group of rare malignant tumors of mesenchymal origin, encompassing over 100 histological subtypes with distinct clinical and molecular characteristics^(1–3)^. Despite advances in molecular profiling, the mechanisms driving sarcomagenesis remain incompletely defined. Deeper investigation into molecular pathways underlying transcriptional dysregulation and epigenetic alterations is needed to uncover therapeutic targets.

ATRX, *α-thalassemia/mental retardation syndrome X-linked*, is a chromatin remodeling protein involved in histone H3.3 deposition, heterochromatin maintenance, and telomere stability^(4)^. A*TRX* loss of function alterations (LOF) have been implicated in various cancers, including gliomas, neuroblastomas, and sarcomas^(5–9)^. In sarcomas, somatic *ATRX* loss has been observed in approximately up to 30% of leiomyosarcomas (LMS) ^(10)^ and undifferentiated pleomorphic sarcomas (UPS)^(11)^, but less common in bone tumors^(12, 13)^. ATRX contains functional domains including the SWI/SNF domain, which modulates chromatin remolding, and the ATRX-DNMT3-DNMT3L (ADD) domain, which regulates transcriptional silencing through DNA methylation^(14, 15)^.

In mesenchymal progenitor cells (MPCs) and UPS models, ATRX loss is linked to epigenetic dysregulation, including de-repression of transposable elements (TEs) and lineage-specific gene programs^(16)^. ATRX loss also activates the alternative lengthening of telomeres (ALT) pathway, associated with poor prognosis in LMS^(17–19)^. TEs influence gene regulation, immunity, and viral mimicry^(20, 21)^; gliomas with ATRX loss display TE and ALT activation, altering the immune microenvironment^(19)^. Sarcoma cell lines with ATRX loss show abnormal interferon signaling^(22)^, suggesting ATRX status may shape immune responses.

ATRX expression has been assessed by immunohistochemistry (IHC) to stratify outcomes, with preserved expression linked to improved survival^(23)^. However, prior datasets were limited by subtype aggregation and insufficient covariate adjustment^(23)^. Thus, the downstream molecular consequences of ATRX loss in sarcomas remain unclear.

Here, we leveraged The Cancer Genome Atlas (TCGA) SARC dataset^(12)^ to assess the molecular and clinical impact of ATRX loss across multiple sarcoma subtypes. Integrative analyses of survival, RNA-sequencing (RNA-seq), and genome-wide DNA methylation revealed that ATRX loss drives widespread DNA hypomethylation, tumor-relevant gene expression changes, and TE de-repression. ATRX loss correlated with significantly shorter survival, particularly in uterine LMS (uLMS), highlighting its potential as prognostic biomarker and opportunities for therapeutic strategies targeting global epigenetic perturbations.

## Methods

We analyzed sarcomas from TCGA Pan-Cancer Atlas SARC cohort^(23)^ focusing on dedifferentiated liposarcoma (DDLPS), UPS, soft tissue LMS (LMS), uterine LMS (uLMS), and myxofibrosarcoma (MFS). These histologies were chosen given their frequency and established prevalence of ATRX alterations (>10%). Patient-level, tumor-level, RNA-sequencing (RNA-seq), and 450K methylation data were accessed via the Genomics Data Portal^(23)^ using TCGAbiolinks^(12)^ and TCGAretriever^(24)^. Additional details for all sections of the methods are provided in the *Extended Methods* in the **Supplemental Materials**.

### Defining ATRX loss

ATRX LOF was defined as nonsense, frameshift, or splice sites mutations, structural deletions, or homozygous copy number losses, consistent with prior criteria^(25)^. In total, 49 sarcomas were classified as ATRX LOF in our dataset (**Table S1**).

### Clinical and tumor covariates

Demographic and clinical variables included age, sex, race/ethnicity, smoking, and radiation exposure. Tumor-level features included histology, anatomic site, mutational burden, and fraction of genome altered. Tumor size was available for 197 cases and categorized as <5 cm, 5-10 cm, or >10 cm. Survival outcomes included overall survival (OS) and disease-free survival (DFS) utilizing vital status and recurrence, respectively, and corresponding time-to-event data from the TCGA.

### Gene expression analysis

RNA-seq data was available for 231 tumors. Preprocessing was conducted using *edgeR*^(26)^, and genes that had low expression across sarcomas were removed (TPM=0 in ≥90% of samples). Normalization was performed using trimmed mean of M-values (TMM)^(27)^, and differential expression was assessed with *limma*-*voom*^(28–30)^. Analyses were adjusted for age, sex, and subtype, and within LMS analyses, analyses included adjustment for uterine vs. non-uterine site. Genes with FDR <0.05 were considered significant^(31)^. Pre-ranked geneset enrichment analyses (GSEA) were performed using clusterProfiler against MSigDB Hallmark gene sets^(32)^. Normalized enrichment scores (NES) and FDR-adjusted p-values were reported. Over-representation of differentially expressed genes was tested using GO Biological Processes with Fisher’s exact tests (FETs), restricted to terms containing 10-500 genes^(27, 33)^.

### DNA methylation

Illumina 450K data were processed with TCGA *SeSAMe* pipeline^(34, 35)^. After exclusion of poor-quality and cross-reactive probes, 289,058 CpGs remained. Beta-values were converted to M-values, and site-specific associations with ATRX loss using *limma*^(28–30)^. Enrichment of CpG and genomic features were assessed with FETs. Differentially methylated regions (DMRs) were identified with *DMR*cate^(36)^. To evaluate cis-regulatory effects, linear models evaluated the associations between CpG methylation and gene expression (transcripts per million [TPM], log2-transformed), and these models were adjusted for age, sex, and histology. Over-representation of significant genes (FDR < 0.05) were then analyzed for functional enrichment among Gene Ontology (GO) biological processes terms to identify pathways impacted by ATRX-associated methylation changes^(37)^.

### Telomere Biology

Telomere length (TL) estimates previously derived by Barthel et al. using whole genome and exome sequencing data from TCGA tumors and the *TelSeq* software*^(^*^32^*^)^* were analyzed as tumor-to-normal TL ratios (log-transformed). Associations with ATRX loss were tested with linear models adjusted for age, sex, histology, and sequencing type. Previously derived telomerase signature scores using RNA-sequencing data from TCGA tumors were analyzed similarly^(38, 39)^. TERRA expression, dichotomized as positive or negative, was compared by ATRX status using FETs^(34, 35)^.

### Immune infiltration

Immune cell proportions from TCGA PanImmune Atlas^(28–30)^ were derived using CIBERSORT^(40)^. Analyses focused on cell types present in ≥10% of tumors. Proportions were compared by ATRX status using Mann-U tests, with stratification by sarcoma subtype.

### Survival analysis

Survival outcomes (OS and DFS) were assessed using Kaplan-Meier (KM) curves and log-rank tests. Cox proportional hazard models estimated hazard ratios (HR) and 95% confidence intervals (CI), adjusting for age, sex, subtype, and tumor size; LMS/uLMS analyses also adjusted for uterine vs. non-uterine site.

Analyses were performed in R 4.3.3. Significance was defined as FDR <0.05 unless otherwise specified.

## Results

### Cohort Characteristics

Our analysis included 234 sarcoma tumors from patients (ages 20-90 years) diagnosed with soft tissue sarcoma subtypes: DDLPS, UPS, LMS, uLMS, and MFS (**Table 1**). Forty-nine (21%) sarcomas had predicted somatic loss of ATRX, consistent with other independent cohorts with somatic alterations or protein expression^(11, 12, 41)^. Among sarcoma subtypes, UPS (n=17/50, 34%) and uLMS (n=9/29, 31%) had the highest frequency of ATRX loss (**Figure 1A**). The distribution of *ATRX* genomic alterations included LOF mutations (n=31, 63%), structural variants (n=3, 6%), and copy number deletions (n=15, 31%) (**Figure 1B, Table S1**). No patient characteristics were associated with *ATRX* status. On average, a larger percentage of genome was altered in sarcomas with ATRX loss vs. retained (mean=45.9 [sd=21.5] and mean=35.1 [sd=25.2], p=0.0036).

**Figure 1.**
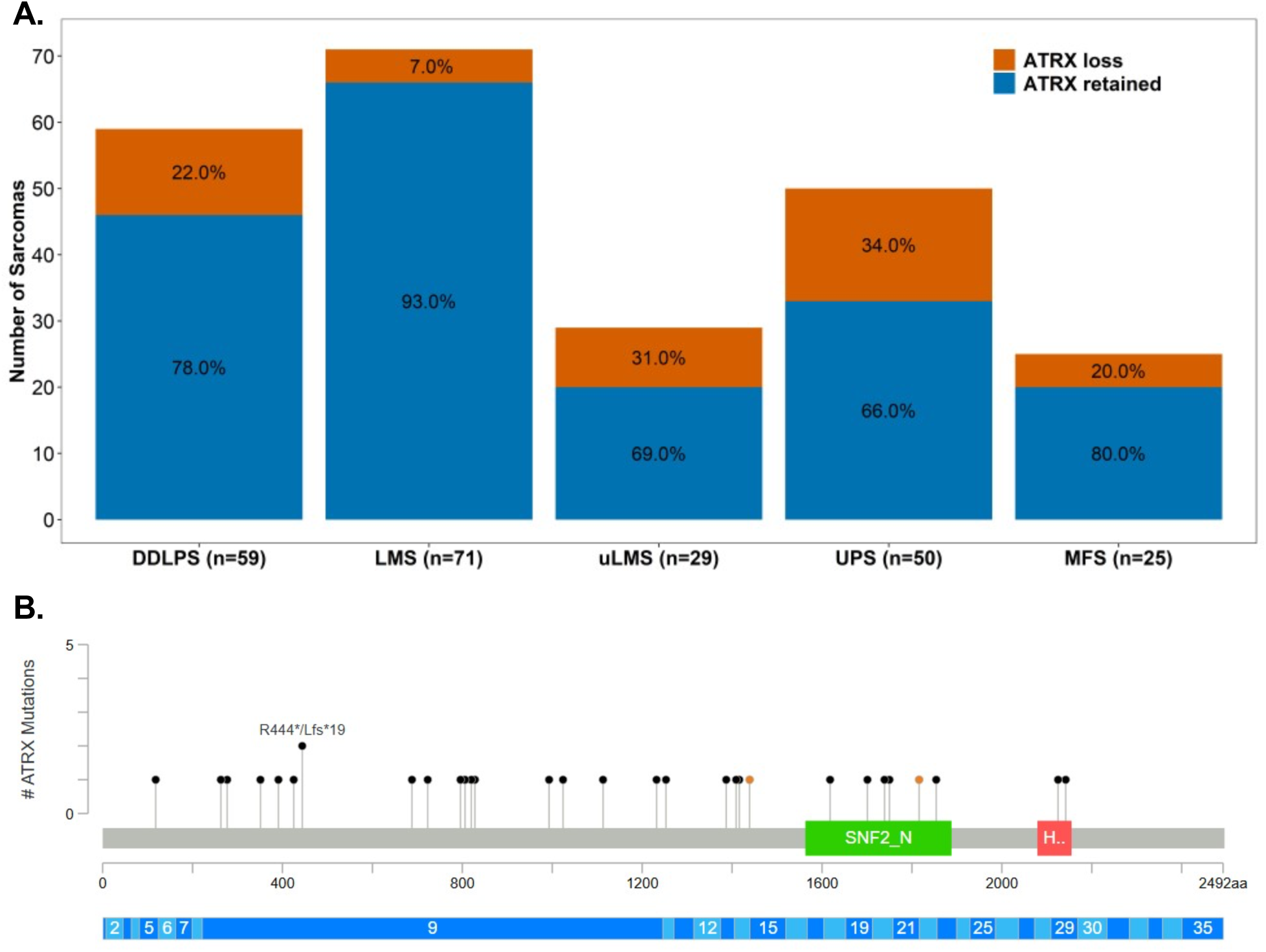
ATRX loss across sarcomas in TCGA. **A)** Distribution of ATRX loss within sarcoma histology subtypes. **B)** Distribution of loss of function (LOF) single variant and frameshift mutations within ATRX gene identified among sarcomas included in analysis (n=31 variants). Plot generated using cBioportal. Variants presented as lollipops, orange and black dots correspond to splice variant and truncating variants. SNF2 family N-terminal (SNF2_N) and Helicase conserved C–terminal domain (Hel) shown in green and red, respectively.

**Table 1.** TCGA study cohort descriptive statistics for sarcomas included in analysis. Distribution reported for all sarcomas included in analysis (overall) and by ATRX status (retained vs. loss). Fisher exact test used to evaluate association between ATRX status and categorical covariates. T-tests used to evaluate association between ATRX status and continuous covariates.

|  | Overall | ATRX retained | ATRX loss | p |
| --- | --- | --- | --- | --- |
|  | 234 | 185 (79%) | 49 (21%) |  |
| Male (%) | 108 (46%) | 82 (44%) | 26 (53%) | 0.23 |
| White (%) | 203 (87%) | 159 (86%) | 44 (90%) | 0.38 |
| Not Hispanic (%) | 201 (86%) | 157 (85%) | 44 (90%) | 0.99 |
| Age (mean [sd]) | 62.5 (13.1) | 62.5 (13.3) | 62.3 (12.3) | 0.89 |
| Cancer Type |  |  |  | 0.0020 |
| Leiomyosarcoma (LMS) | 71 (30%) | 66 (36%) | 5 (10%) |  |
| Uterine Leiomyosarcoma (uLMS) | 29 (12%) | 20 (11%) | 9 (18%) |  |
| Dedifferentiated Liposarcoma (DDLPS) | 59 (25%) | 46 (25%) | 13 (27%) |  |
| Undifferentiated Pleomorphic Sarcoma (UPS) | 50 (21%) | 33 (18%) | 17 (35%) |  |
| Myxofibrosarcoma (MFS) | 25 (11%) | 20 (11%) | 5 (10%) |  |
| Mutational Count (geometric mean [sd]) | 44.9 (2.2) | 43.2 (2.3) | 51.6 (1.9) | 0.13 |
| Fraction of Genome Altered (expressed as percentage) | 37.3 (25.2) | 35.1 (25.2) | 45.9 (21.5) | 0.0036 |
| Radiation Treatment (among reported) |  |  |  | 0.0051 |
| Yes | 58 (26%) | 38 (22%) | 20 (43%) |  |
| No | 165 (74%) | 138 (78%) | 27 (57%) |  |
| Tumor Size (mean [sd]) | 13.0 (8.0) | 13.0 (8.3) | 13.0 (6.6) | 0.99 |
| Tumor Size |  |  |  | 0.52 |
| < 5 cm | 22 (9%) | 18 (10%) | 4 (8%) |  |
| 5-10cm | 65 (28%) | 55 (30%) | 10 (20%) |  |
| 10+ cm | 110 (47%) | 85 (46%) | 25 (51%) |  |
| Missing | 37 (16%) | 27 (15%) | 10 (20%) |  |

### ATRX loss in sarcomas is associated with subtype-specific transcriptional dysregulation

ATRX maintains heterochromatin structure and regulates gene expression^(42, 43)^. Given prior evidence of transcriptional dysregulation in ATRX-deficient MPCs^(16)^, we evaluated transcriptome-wide changes in sarcomas. ATRX loss was associated with reduced *ATRX* expression across all subtypes (**Figure 2A**, p_wilcox_ < 0.05). Adjusted analyses identified 27 differentially expressed genes (DEGs) overall (FDR < 0.05), with *ATRX* as the most significant association observed (log_2_-FC= −1.08 [95% CI: −1.41, −0.75], p=9.2×10^-10^, FDR=1.9×10^-5^) (**Figure S1, Table S2, Table S3, Data Table 1**).

**Figure 2.**
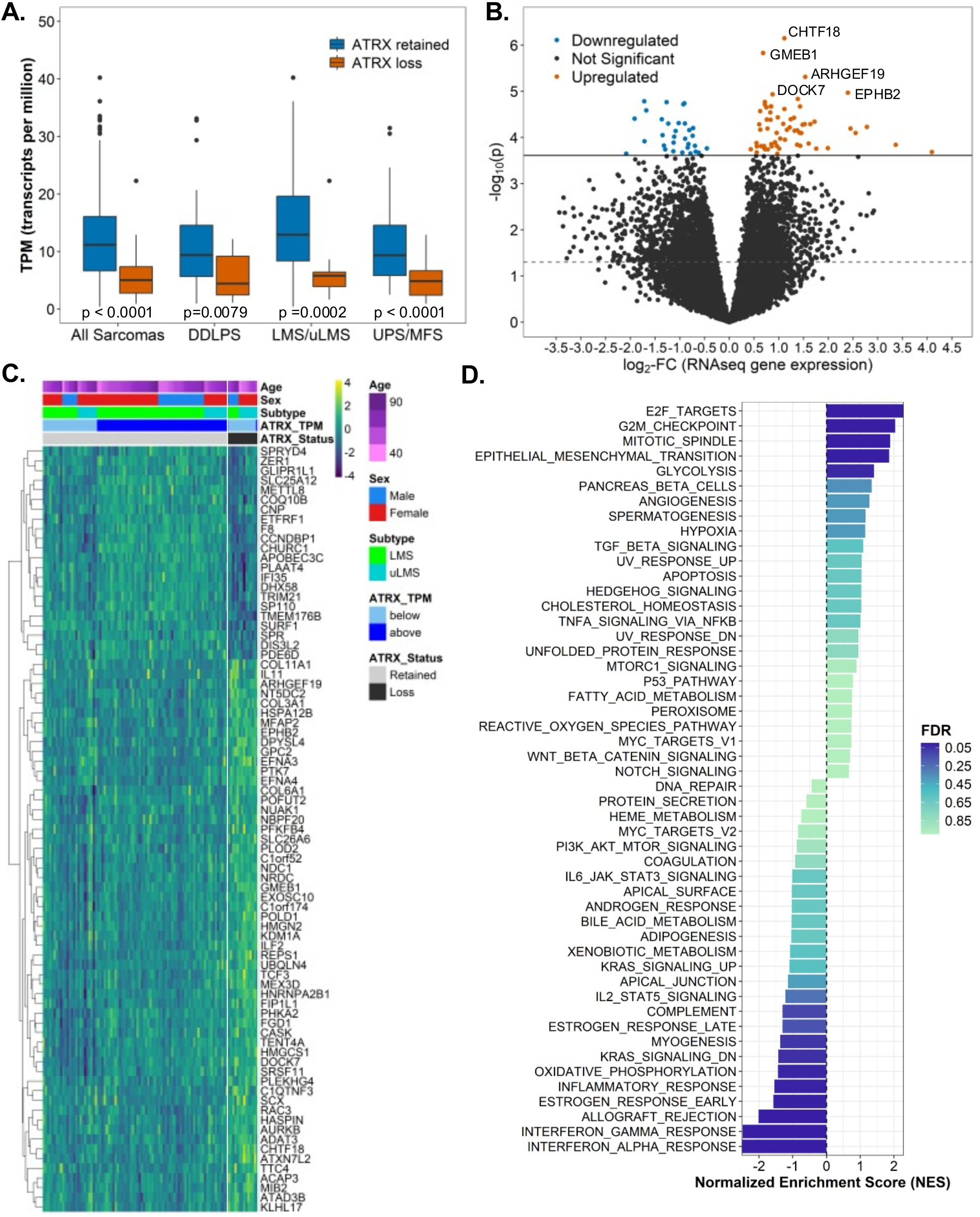
Impact of ATRX loss on gene expression in sarcomas. **A**) Boxplot presents distribution of ATRX gene expression (transcripts per million [TPM] among all sarcomas and stratified by histologic subtype. ATRX retained and loss is shown in blue and red, respectively. All data shown except for one outlier, where TPM=99 for a DDPLS tumor. P-values obtained from Wilcoxon-Mann U test. **B**) Volcano plot presents log_2_-fold change (log_2_-FC) estimates and –log_10_-transformed p-values across all gene expressed in LMS/uLMS. Estimates and p-values obtained from adjusted linear model examining the impact of ATRX loss on normalized gene counts. Models adjusted for tumor site (uterine, non-uterine), age at diagnosis, and sex. Blue, red, and dark gray points correspond to gene expression changes that were downregulated in LMS with ATRX loss (FDR < 0.05, log_2_-FC < 0), upregulated in LMS with ATRX loss (FDR < 0.05, log_2_-FC > 0), and not associated with ATRX loss (FDR > 0.05), respectively. Solid and dotted line correspond to FDR < 0.05 threshold and p < 0.05, respectively. **C**) Heatmap of gene expression (protein-coding only) associated with ATRX loss (FDR < 0.05, n=79 genes). Scaled, log_2_-transformed TPM values presented in plots and gene clusters (i.e., row clusters) identified using consensus clustering (see Methods for more details). **D**) Normalized enrichment scores (NES) across hallmark gene sets (see Methods). NES generated using GSEA approach and ranked gene list by log_2_-FC. NES plotted by size and direction and shaded by FDR of enrichment score p-value. Gene sets with NES < 0 and NES > 0 correspond to sets that are downregulated and upregulated by ATRX loss, respectively.

Subtype-specific analyses revealed that no DEGs were identified with ATRX loss in DDLPS (FDR > 0.05), but 94 and 17 DEGs with ATRX loss in LMS/uLMS and UPS/MFS were identified (FDR < 0.05), respectively. Most DEGs in UPS/MFS (15/17) were upregulated with ATRX loss, including *SS18L1* (implicated in SWI/SNF dysregulation), *WNK3* (cell survival), and *MSI1* (stem cell marker) (**Table S4**)^(12)^. In LMS/uLMS, 63 DEGs were upregulated with ATRX loss, and these DEGs mapped to telomere maintenance (*HNRNPA2B1, AURKB, EXOSC10*), Wnt signaling (*ARHGEF19, PTK7*), and p53 signaling (*NUAK1, AURKB, KDM1A*) pathways; downregulated DEGs notably mapped to innate immunity (*DHX58, IFI35, TRIM21, APOBEC3C*) pathways (**Figure 2B, Table S5, Data Table 2**).

Among the Hallmark genesets, GSEA identified global upregulation of G2M checkpoint and epithelial-mesenchymal transition pathways, and downregulation of interferon and IL6-JAK-STAT signaling with ATRX loss across all sarcoma types (FDR <0.05) (**Table S6**, **Figure S1, Figure S2**). ATRX loss in LMS/uLMS tumors was associated with upregulation of MYC targets and unfolded protein response pathways while it was associated with downregulation of MYC targets in UPS/MFS, illustrating distinct subtype-specific transcriptional effects of ATRX loss (**Figure 2D, Table S6**).

### Sarcomas with ATRX loss are characterized by genome-wide DNA hypomethylation

ATRX-deficient sarcomas were characterized by widespread DNA hypomethylation *in vitro*^(14)^. Given ATRX’s role in maintaining chromatin states through DNA methylation^(44)^, we examined methylation changes across the genome. We identified 269 CpGs differentially methylated by ATRX status (FDR < 0.05) (**Figure 3A**); 87% were associated with decreased methylation, indicating a preferential hypomethylating impact (**Figure 3B**, **Data Table 3**). ATRX-associated CpGs were enriched in CpG shores (p_FET_ < 0.05), regions where methylation is less stringently maintained during DNA replication (**Figure S3**). Differentially methylated CpGs annotated to genes, including *BMF*, *DRG2*, and *CCNI* which regulate apoptosis, cell cycle progression, and proliferation, respectively^(45–47)^. We identified 16 DMRs, including a region of hypomethylation within the DAXX gene body, previously known to complex with ATRX, and a region of hypermethylation in DRG2, previously linked to downregulation in ATRX-deficient sarcomas and ALT-associated tumors (**Data Table 4, Table S3**)^(48)^.

**Figure 3.**
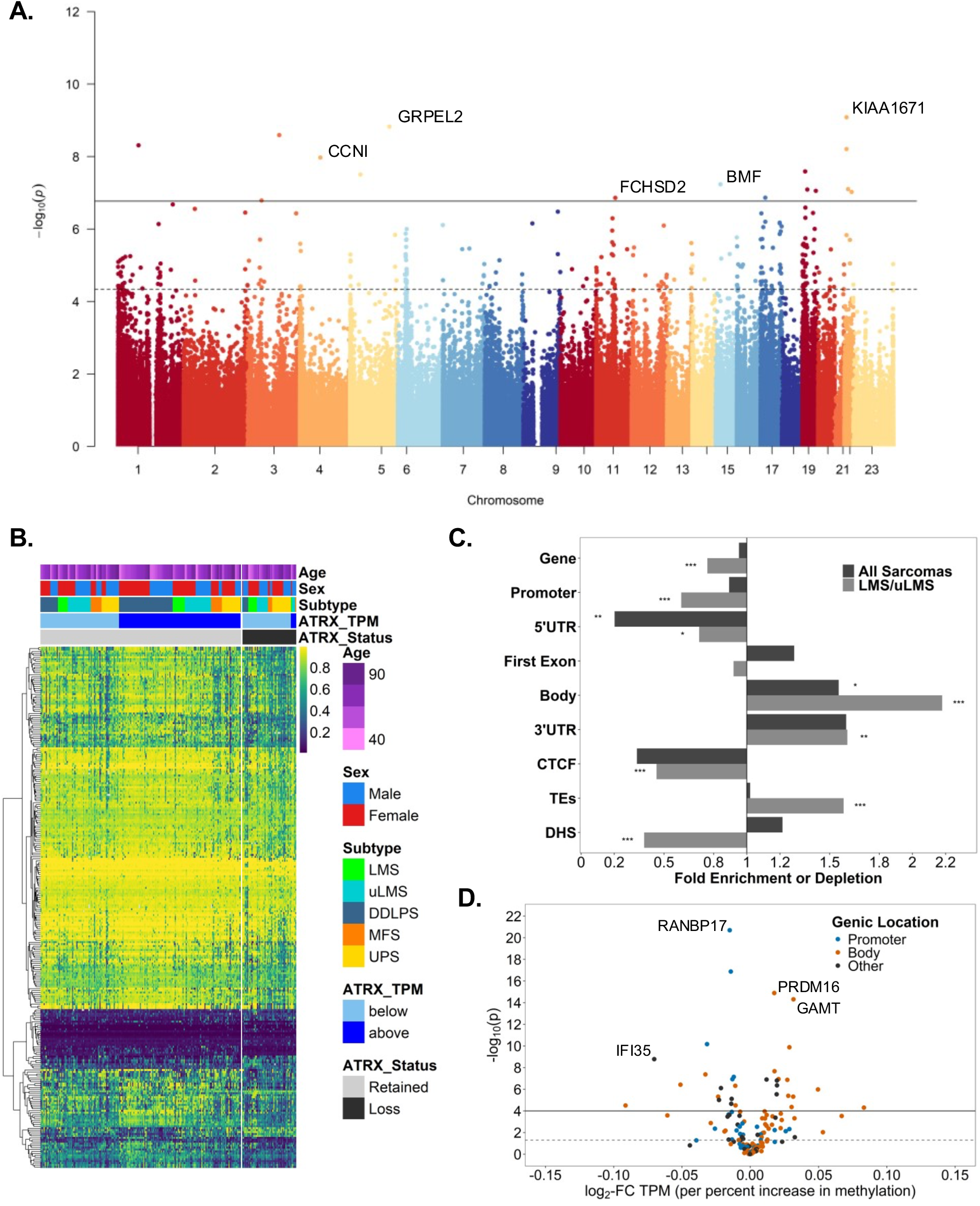
Impact of ATRX loss on genome-wide DNA methylation across sarcoma subtypes in the TCGA. **A**) Manhattan plot of –log10-transformed p-values from associations between ATRX loss and CpG-specific methylation (M-values) from linear model adjusted for age, sex, and histology subtype using limma (see Methods for more details). Solid and dashed lines correspond to Bonferroni adjusted _p_-value threshold ^(p=1.7×10-7)^ and FDR=0.05 threshold ^(p=4.6×10-5)^, respectively. **B**) Heatmap of CpGs associated with ATRX loss (FDR < 0.05, n=269). Beta-values (range: 0 to 1) are presented in plots and gene clusters (e.g., row clusters) identified using consensus clustering (see Methods for more details). **C**) Enrichment of ATRX loss–associated methylation among genomic features (FDR < 0.05). Odds ratios presented for each feature and p-values obtained from Fisher Exact Tests. Features evaluated included gene, promoter (within 200bp and 1500bp of gene start), 5’ untranslated region (UTR’5), first exon, gene body (body), 3’ untranslated region (UTR’3), CCCTC-binding transcription factor (CTCF), transposable element region (TES), and DNase hypersensitivity region (DHS). **D**) Association between ATRX-associated methylation and cis-gene expression. Association estimates from linear models of ATRX-associated methylation on log_2_-transformed TPM expression adjusted for age, sex, and histologic subtype. Log_2_-fold changes (Log_2_-FC) presented on x-axis and – log_10_-transformed p-values presented on y-axis. Associations were characterized by the location of the ATRX-associated methylation.

Subtype analysis revealed no associations between CpG-specific methylation and ATRX loss in DDLPS and UPS/MFS (FDR > 0.05); however, ATRX loss was strongly associated with differential methylation in LMS/uLMS (2243 CpGs, FDR < 0.05) (**Figure S4, Data Table 5**), with 91% of differentially methylated CpGs associated with decreased methylation. ATRX-associated CpGs were enriched in TE regions, consistent with hypomethylation-driven TE derepression^(49)^, but depleted in genic regions, CCCCTC-Binding factor (CTCF) binding, and DNase hypersensitive sites (DHS) (p_FET_ < 0.05), suggesting ATRX loss preferentially disrupts methylation within repetitive elements (e.g., TE) rather than chromatin accessibility loci (e.g., DHS, CTCF) (**Figure 3C**). We identified 363 DMRs in LMS/uLMS, including in *DAXX* and fibroblast growth factor genes (*FGF6, FGF23, FGF3, FGF21*) (**Data Table 6**).

Among the differentially methylated CpGs associated with ATRX loss that annotated to a gene, 149 (122 unique genes) and 1138 (643 unique genes) CpGs corresponded to a gene with expression across all sarcomas and LMS/uLMS, respectively. Methylation at 74 and 653 CpGs was associated with corresponding gene expression among all sarcomas (**Data Table 7**) and within LMS/uLMS (**Data Table 8**). These CpGs were more often positively correlated with expression (52.7% in all sarcomas; 85.6% in LMS/uLMS) and located in gene bodies (54.1% and 69.5%, respectively). Pathway analyses demonstrated enrichment in MHC protein complex assembly and RAS signaling across all sarcomas, while LMS/uLMS additionally showed enrichment in reactive oxygen species (ROS)-related metabolic processes (**Data Tables 9-10**).

### Expression of TEs is enhanced in sarcomas with ATRX loss

Among 109 sarcomas with TE expression data^(50)^, 96 TEs were differentially expressed with ATRX loss in sarcomas (p < 0.05), with 97% (n=93) upregulated, underscoring ATRX’s role in TE silencing (**Figure 4A, Data Table 11**). TEs associated with ATRX loss were enriched in LINE and LTR classes and in ERV and L1 fin long interspersed nuclear elements (LINE) and long terminal repeat (LTR) classes and ERV and L1 groups (p_FET_ < 0.05)(**Figure 4B-4C**), consistent with prior *Atrx* KO MPC studies showing de-repression of ERV and LINE-1 elements^(16)^.

**Figure 4.**
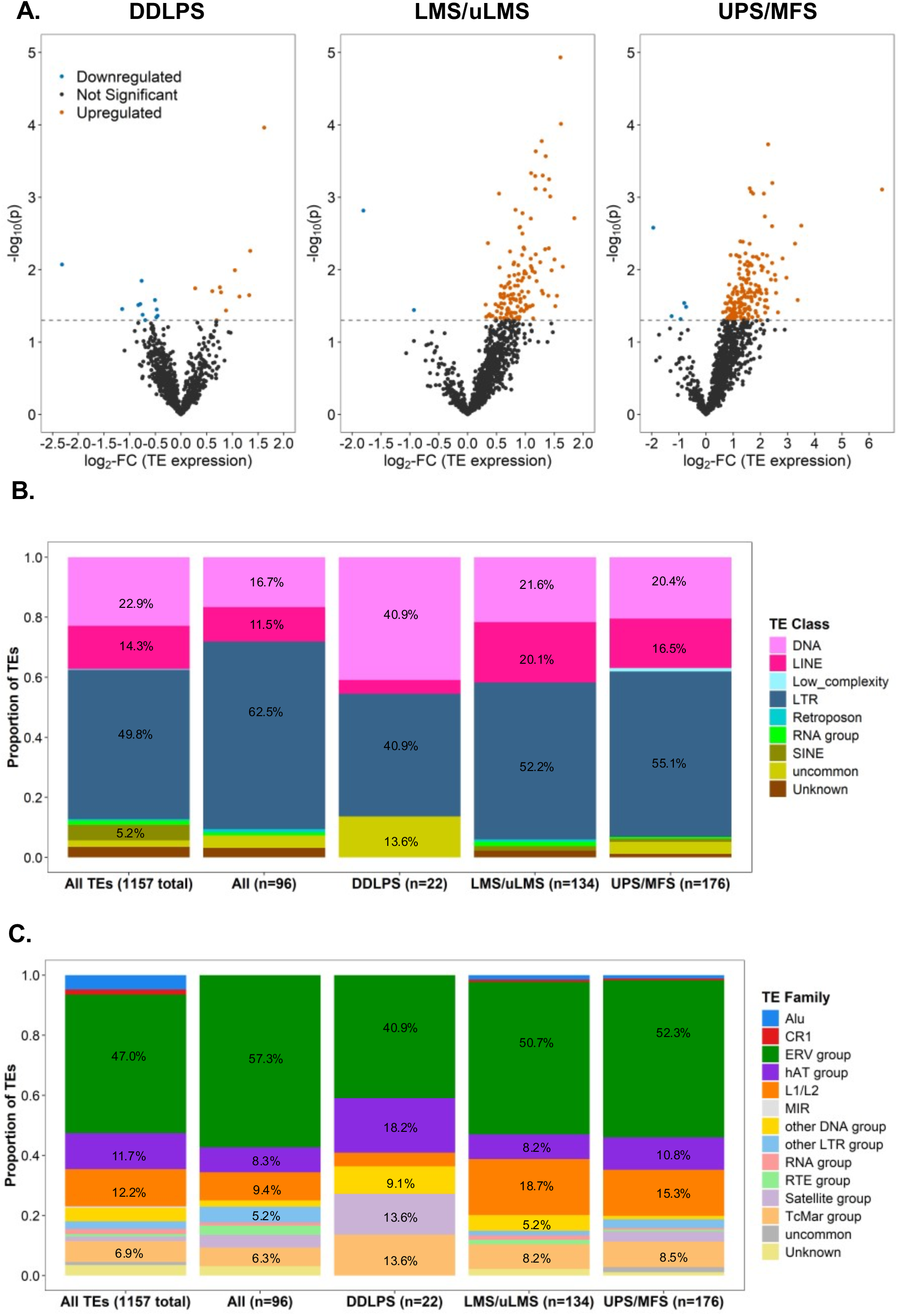
Impact of ATRX loss on transposable element (TE) expression across sarcoma subtypes in the TCGA. **A**) Associations between TE expression and ATRX loss within each histologic subtype. Log_2_-fold change (Log_2_-FC) and –log_10_(p-value) from linear models evaluating ATRX loss on TE expression using limma (see Methods for more details). Models adjusted for age, sex, and histologic subtype (for LMS/uLMS and UPS/MFS). Blue, red, and dark gray points correspond to TE expression changes that were downregulated with ATRX loss (FDR < 0.05, log_2_-FC < 0), upregulated in LMS with ATRX loss (FDR < 0.05, log_2_-FC > 0), and not associated with ATRX loss (FDR > 0.05), respectively. **B**) Distribution of ATRX-associated TE expression (p < 0.05) by TE sequence class overall and by histologic subtype. A total of 1157 TEs had detectable expression across sarcomas, and n corresponds to the number of TEs associated with ATRX loss at a p-value < 0.05 threshold. Proportion of TEs within each class are shown for those that account for more than 0.05 (or 5%). **C**) Distribution of ATRX-associated TE expression (p < 0.05) by sequence family overall and by histologic subtype. See information for **B)** for more detail

TE dysregulation was most pronounced in LMS/uLMS and UPS/MFS with ATRX loss, where 134 and 176 TEs were differentially expressed, respectively, with >99% upregulated in sarcomas with ATRX loss. In UPS/MFS, TEs associated with ATRX loss were significantly enriched in LTR elements (p_FET_=0.013), whereas TEs associated with ATRX loss that were identified in LMS/uLMS were enriched in both LTR and LINE classes (p_FET_=0.052). At the TE family level, UPS/MFS tumors displayed a distinct overrepresentation of ERV and LINE1/LINE2 families (p_FET_ < 0.001) (**Figures 4B-4C**).

### ATRX loss impacts immune infiltration in sarcomas

Using immune infiltrate estimates derived from Cibersort^(40, 51)^, we evaluated the association between ATRX status and proportion of immune cell types (**Figure 5A** and **Table S7**). Compared to sarcomas with ATRX retained, sarcomas with ATRX loss had lower median proportion of CD4+T cells (0.12 vs. 0.09, p_wilcox_=0.032), specifically CD4+T resting memory cells (0.09 vs. 0.06, p_wilcox_=0.046), and higher median proportion of M2 macrophages (0.39 vs. 0.47, p_wilcox_=0.018). The reduction in resting memory CD4+ T cells associated with ATRX loss may reflect impaired immune readiness and contribute to the immune-suppressive tumor microenvironment. Among subtypes, DDLPS with ATRX loss had lower median proportion of CD4+T cells (0.13 vs. 0.08, p_wilcox_=0.0074). LMS/uLMS sarcomas with ATRX loss (when compared to ATRX retained) had lower B-cells (0.03 vs. 0.01, p_wilcox_=0.0065) and higher macrophages M0 (0.00 vs. 0.09, p_wilcox_=0.0015). The median proportion of regulatory T-cells was lower in UPS/MPS with ATRX loss vs. retained (0.02 vs. 0.01, p_wilcox_=0.015).

**Figure 5.**
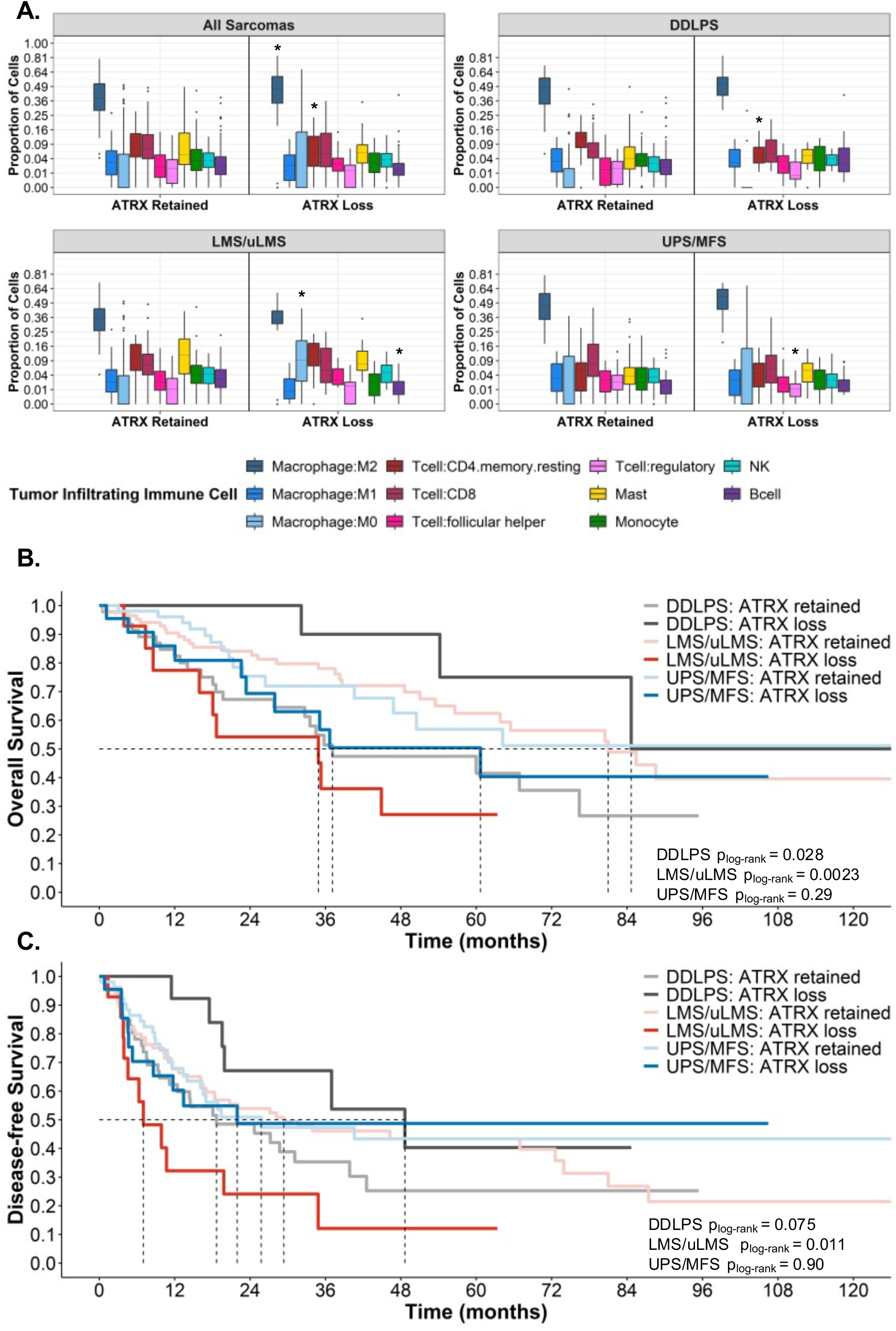
ATRX loss has prognostic implications related to tumor immune infiltration and survival among subtypes. **A**) Distribution of the proportion of tumor infiltrating immune cells across all subtypes and within subtypes. Immune cell proportions estimated from gene expression data using Cibersort (see Methods for more details). Mann-U tests compared distribution of cell proportions by ATRX status, and * denote p-value < 0.05. For full results, see Table S7. **B**) Kaplan-Meier curves presented for overall survival within histologic subtypes and stratified by ATRX status. Lighter and darker curves correspond to ATRX retained and loss, respectively. Gray, red, and blue lines correspond to DDLPS, LMS/uLMS, and UPS/MFS, respectively. Dotted lines correspond to median survival (in months). P-values from log-rank tests comparing survival distribution between ATRX retained vs. loss within each histology subtype. For additional results see Table S8. Results for uLMS, LMS, UPS, and MFS provided in Figure S6. **C**) Kaplan-Meier curves presented for disease-free survival within histologic subtypes and stratified by ATRX status. See **B)** for more information.

### miRNA expression is dysregulated in sarcomas with ATRX loss

ATRX loss broadly impacts miRNA regulation. Expression of 34 miRNAs was upregulated in sarcomas with ATRX loss while expression of three miRNAs were downregulated, including hsa-mir-34a, hsa-mir-34c, and hsa-mir-150 (p < 0.05), consistent with the role of *ATRX* as an epigenetic silencer (**Figure S5, Data Table 12**). Both hsa-mir-296 and hsa-mir-197 reached an FDR=0.05, and these miRNAs have been shown to associated with angiogenesis, a critical process for tumor growth and metastasis^(52, 53)^. When stratified by subtype, ATRX loss was associated with miRNA expression for five, 27 (79.3% upregulated), and 29 (92.6% upregulated) transcripts in DDLPS, LMS/uLMS, and UPS/MFS, respectively (p < 0.05). Unique to LMS/uLMS, let-7e, let-7f-1, and let-7f-2 expression was increased in LMS with ATRX loss compared to those with ATRX retained (FDR < 0.05), potentially consistent with prior analysis of LMS TCGA samples^(13)^, where miRNA expression patterns had distinctly lower expression in LMS/uLMS.

### Telomere length is significantly longer in tumors with ATRX loss compared to those with ATRX retained

Prior studies have shown that *ATRX* or *DAXX* mutations promote ALT-mediated telomere elongation, resulting in longer telomeres than in wild-type tumors^(10, 54)^. In our analysis of the tumor to normal TL ratio, sarcomas with ATRX loss had significantly longer TL relative to normal tissue compared to ATRX-retained cases (β=0.29 [se=0.14], p=0.036), after adjustment for age, sex, subtype, and sequencing type (**Figure S6**). This effect was strongest among UPS/MFS (β=0.77 [se=0.21], p=0.0058). Telomerase expression scores were not associated with ATRX status. Notably, ATRX loss conferred a 2.70-fold (95% CI: 1.00, 9.32) higher likelihood of positive TERRA expression (p_FET_=0.046); all UPS/MFS with ATRX loss expressed TERRA (p_FET_=0.029). These findings align with prior evidence that TERRA at telomeres promotes instability and ALT-mediated telomere elongation in sarcomas^(55)^.

### ATRX loss correlates with poor overall survival (OS) in leiomyosarcoma

ATRX loss was associated with shorter OS in LMS/uLMS (median survival of 81.0 vs 34.9 months for ATRX retained vs. loss [p_log-rank_=0.0023]) (**Figure 5B, Table S8**). This association persisted after adjustment for age, sex, tumor size, and subtype (LMS/uLMS) (HR=2.89 [95% CI: 1.11, 7.49], p=0.030). Within LMS/uLMS, ATRX loss was associated with shorter DFS (median survival of 28.2 vs. 7.0 months, for ATRX retained vs. loss, respectively, p_log-rank_ =0.011) (**Figure 5C**). However, this association was attenuated after adjustment for tumor size (p_cox-PH_=0.22). The negative correlation with OS persisted in the uLMS cohort and no effect of ATRX status on DFS in LMS or uLMS subtypes after adjusting for age and tumor size (**Figure S7**). These findings suggest that ATRX loss may serve as a prognostic indicator for worse OS in patients with uLMS.

ATRX loss exerted a modest protective effect on OS and DFS in DDLPS. ATRX loss was associated with longer OS in DDLPS (median survival of 84.7 vs 37.1 months for ATRX loss and retained [p_log-rank_=0.028]). However, this association was attenuated in the Cox-proportional hazards model after adjusting for tumor size, age, and sex (HR=0.25 [95% CI: 0.06, 1.11], p=0.068). DFS was also longer in DDLPS with ATRX loss, median DFS 48.7 months, compared to those with ATRX retained, median DFS 18.7 months (p_log-rank_=0.075). This association persisted after adjusting for tumor size, age, and sex (HR=0.31 [95% CI: 0.10, 0.93], p=0.036).

## Discussion

Soft tissues sarcomas exhibit ATRX loss in up to 30% of cases; however, the biologic consequences and therapeutic implications remain unclear. Here, we profiled the molecular and clinical landscape of ATRX loss across sarcoma subtypes using genomic, transcriptomic, and epigenomic data from TCGA. ATRX loss occurred in approximately 21% of cases, most frequently in UPS and uLMS, and was associated with subtype-specific transcriptional dysregulation, genome-wide DNA hypomethylation, and derepression of TEs, particularly in LMS/uLMS and UPS/MFS. Tumors with ATRX loss demonstrated altered immune infiltration with reduced proportion of CD4+ T cells and increased proportion of M2 macrophages. Clinically, ATRX loss predicted worse OS in LMS/uLMS, but favorable outcomes in DDLPS, suggesting its context-dependent role in sarcoma tumor biology.

ATRX loss produced widespread transcriptional dysregulation, particularly of genes linked to tumor progression. TE expression was markedly increased, consistent with ATRX’s known role in silencing TEs in MPCs^(16)^. Widespread DNA hypomethylation accompanied ATRX loss, likely contributing to transcriptional activation of genes and TEs. These findings emphasize the important role of ATRX in transcriptional silencing highlighting their potential utility in future mechanistic studies.

Mechanistically, ATRX-deficient MPCs show increased chromatin accessibility, including the Wnt-β-catenin signaling genes, which acquire active chromatin marks in ATRX-deficient UPS^(16)^. Consistent with that observation, WNT-β-catenin signaling genes were transcriptionally upregulated, suggesting ATRX loss could promote tumor plasticity and resistance to differentiation. Prior work showed that UPS cells with ATRX loss were more sensitive to tegavivint, a β-catenin antagonist, supporting WNT dependency in this context^(56)^. Given that tegavivint is in clinical trials for other mesenchymal tumors (NCT04851119), WNT-targeted therapy merits exploration in ATRX-deficient sarcomas.

ATRX loss was associated with epigenetic dysregulation in the form of genome-wide hypomethylating effect. Several differentially methylated genes, including *BMF*, *DRG2*, and *CCNI*, regulate apoptosis, cell cycle progression, and proliferation^(16)^. Moreover, we observed downregulation of interferon signaling, consistent with prior UPS and MPC models^(16)^, suggesting a compensatory mechanism to suppress innate immune activation downstream of TE expression. In LMS, ATRX loss correlated with decreased expression of innate immune regulators such as *DHX58 and TRIM21*. Their loss may blunt innate immune responses, fostering an immune-suppressive tumor microenvironment. These findings highlight the interplay between ATRX loss, TE depression, and immune modulation, suggesting therapeutic opportunities to restore innate immune signaling and enhance immunotherapy responsiveness.

Survival analysis revealed striking subtype-specific differences. In LMS/uLMS, ATRX loss was associated with significantly worse OS and DFS, particularly in uLMS, where ATRX loss is more prevalent. Whether this effect is absent in LMS due to distinct biology or limited sample size remains uncertain. In contrast, ATRX loss in DDLPS correlated with improved survival and lacked major transcriptional or methylation effects, underscoring subtype heterogeneity in ATRX function. Together, this data suggests ATRX loss serves as a context-dependent modifier of sarcoma biology, with divergent effects across subtypes.

While our findings provide valuable insight, it is important to acknowledge its limitations. Sample sizes were modest, reflecting the rarity of sarcoma, and require validation in larger cohorts. Heterochromatin profiling was not feasible in tumor samples, limiting integration with prior cell line studies. Methylation data were derived from 450K arrays rather than whole-genome profiling, which may restrict resolution of ATRX-associated methylation changes. Finally, single-cell RNA-seq could clarify how ATRX loss shapes immune cell populations within the tumor microenvironment.

Overall, ATRX loss impacts sarcoma biology through transcriptional dysregulation, DNA hypomethylation, TE depression, and modulation of immune signaling, with distinct consequences across subtypes. In LMS/uLMS, ATRX loss portends poor outcomes and highlights potential vulnerabilities in WNT signaling, ALT-mediated telomere maintenance, and innate immune suppression. Conversely in DDLPS, ATRX loss associated with improved survival, emphasizing the need for subtype-specific analyses. These results not only advance understanding of ATRX biology in sarcomas but also highlight therapeutic opportunities including epigenetic modulators, WNT-β-catenin inhibitors, ALT-targeted approaches, and strategies to enhance immunogenicity. Future studies should investigate ATRX as a clinically actionable biomarker to refine treatment strategies in sarcoma.

## Data Availability

The study only used data that is available from the TCGA database and published manuscripts as cited.

## Acknowledgements

We gratefully acknowledge the support of the Burroughs Wellcome Fund/Physician Scientist Incubator Program and the T32 Award (5T32HD071834) for Research Training for Pediatric Subspecialty Fellows through the Department of Pediatrics at UPMC Children’s Hospital of Pittsburgh. B.A.N. is a Damon Runyon Clinical Investigator, supported in part by the Damon Runyon Cancer Foundation (CI-124-23) as well as NCI K08 CA245212. The Hillman Cancer Center Support Grant (P30 CA047904) partly supported this work through shared resource centers. This project used the UPMC Hillman Cancer Center (HCC) Cancer Biostatistics Facility.

## Supplemental Materials

### Methods

#### Sample selection from TCGA

We included sarcomas within the SARC study in the Pan-Cancer Atlas initiative leveraging samples and data from TCGA^(1)^. Corresponding patient-level, tumor-level, RNA-sequencing, and 450K methylation data were available for each sample from the Genomics Data Portal^(2)^. Data were accessed using R packages TCGAbiolinks^(3–5)^ and TCGAretriever^(6)^. The following sarcoma subtypes were included: DDLPS, LMS, uLMS, UPS, MFS. ICD-O-10 codes were used to identify uLMS (C54.X or C55). These subtypes were included as they are common histologic subtypes of sarcomas and known to frequently harbor *ATRX* alterations (>10% of tumors within each subtype have ATRX loss).

#### Defining somatic ATRX loss

We determined somatic *ATRX* loss of function (LOF) using somatic single variant, structural variant, and copy number variation data. *ATRX* status was determined based on the presence or absence of genetic lesions predicted to confer a LOF phenotype based on prior work (e.g., nonsense or frameshift mutations or deep deletions)^(7, 8)^. Somatic mutations were included if the variant annotation corresponded to frameshift insertion or deletion, nonsense mutation, or splice variant. We also included sarcomas with structural deletions. Finally, we assigned sarcomas with copy number homozygous deletions of *ATRX* as ATRX-deficient. We expected that these mutations would result in pathogenic loss of ATRX. We identified 49 sarcomas with ATRX LOF in our dataset (**Table S1**).

#### Participant and tumor-level covariates

We extracted the following patient-level covariates: age at diagnosis, sex, race/ethnicity, smoking status, and receipt of radiation therapy. The following tumor-related variables were downloaded or provided by TCGA, including cancer site, histology (i.e., subtype), mutational count, and fraction of genome altered. Tumor size variable was initially available ∼75% of cases, and we manually reviewed additional pathologic reports available via TCGA portal to ascertain tumor size for 18 additional sarcomas^(9)^. A total of 197 sarcomas had tumor size information. This tumor size variable was categorized as ≥ 10 cm, 5-10 cm, and < 5 cm. For all tumors we extracted vital status (alive/dead), recurrence (yes/no), and corresponding follow-up time for these outcomes. Corresponding survival time data was provided by TCGA and expressed in months.

#### Gene expression

RNA-sequencing gene expression data was downloaded and available for 231 tumors included in our analyses. We preprocessed our RNA-sequencing data using *edgeR*^(10, 11)^. Using transcripts per million (TPM) data, we removed genes with no expression (TPM=0) in 90% or more of samples. We then removed these genes from the RNA-seq data and calculated normalization factors using the trimmed mean method (TMM)^(12)^. All analyses were conducted in *limma* with the *voom* transformation^(13–15)^ to evaluate the impact of ATRX loss on gene expression. We adjusted for age, sex, and histology subtype (when relevant) in all models. For analyses of LMS, we adjusted for cancer site (non-uterine vs. uterine). The empirical Bayes moderated t-statistics were extracted using the *eBayes* function in *limma*. Genes were considered differentially expressed if they reached a false discovery rate (FDR) < 0.05^(16)^. All association estimates were expressed in log_2_-fold change (log_2_-FC) and corresponding 95% confidence intervals (CI) were extracted. Clustering analyses were conducted on genes differentially expressed in sarcomas with ATRX loss. Hierarchical clustering on the genes (rows) was performed using Pearson correlation (1-r) for distance and complete linkage.

#### Geneset analyses

Using the clusterProfiler package in R^(17)^, we conducted geneset enrichment analyses (GSEA) among the Hallmark gene sets^(18)^ using pre-ranked gene lists ordered by log_2_-FC. Normalized enrichment scores (NES) were extracted from results (to account for gene set size), and p-value (p) obtained from permutation-tests and adjusted p-value using Benjamini-Hochberg adjustment (FDR). The percentage of genes (%) that contribute to the enrichment score are determined based on the peak of the enrichment score. We also conducted over-representation analyses of the differentially expressed genes (FDR < 0.05) in LMS/uLMS among Gene Ontology (GO) biologic processes terms^(19, 20)^ by the direction of association (upregulated, downregulated) using enrichGO function. Significance of the tests were determined from Fisher’s Exact Tests (FET). Terms were analyzed if they contained between 10-500 genes. We reported all terms that reached a nominal p-value threshold of 0.05.

#### Genome-wide DNA methylation

Illumina DNA methylation data 450K was obtained for all tumors. The data was previously preprocessed using TCGA Methylation Array Harmonization Workflow with the SeSAMe pipeline ^(21, 22)^. Using the SeSAMe methylation beta-value data, we filtered CpG probes with more than 5% missing values across samples, those annotating to a known SNP, Y-chromosome, and cross-reactive probes. We retained 289,058 CpGs for our analysis. Beta-values were converted to M-Values (log_2_[beta-values/ [1-beta-values]]). We examined the impact of ATRX loss on site-specific methylation using *limma*^(13–15)^. We evaluated the distribution of relation to CpG between ATRX-associated CpGs and non-ATRX associated CpGs using FET. The enrichment or depletion of genomic features between ATRX-associated CpGs and non-ATRX associated CpGs was evaluated using FET and extracted corresponding odds ratios (OR) and 95% CI. Differentially methylated region (DMR) analyses to examine regions of DNA methylation associated with ATRX loss using DMRcate (using lambda=1000, C=2)^(23)^.

We further evaluated the functional impact of *ATRX*-associated methylation on *cis* gene expression using log_2_-transformed TPM data among all sarcomas and the LMS/uLMS group. Among the *ATRX*-associated CpGs that annotated to a gene, linear models were employed to evaluate the association between methylation (beta-value data) and expression (using log_2_-transformed TPM data). We adjusted for age, sex, and histology subtype in models with all sarcomas. For analyses of LMS/uLMS, we adjusted for anatomic site (i.e., non-uterine vs. uterine). Among the genes with *cis* methylation-expression associations (p < 0.05), we conducted over-representation analyses using *enrichGO* function in clusterProfiler and examined the over-representation of genes in Gene Ontology Biologic Processes terms^(19, 20)^. Terms were analyzed if they contained between 10-500 genes and reported if reached a nominal p-value threshold of 0.05.

#### Transposable element expression

Using the data from a previous study by Kong et al. examining TE expression across TCGA samples^(24)^, we extracted TE expression data from https://github.com/ucsffrancislab/REdiscoverTE/tree/master/original/REdiscoverTEdata/inst/Fig4_data.

In total, 109 sarcoma samples had available TE expression data. TE expression values were reported in TPM and corresponded to 1206 TEs. We utilized the annotation file from the atena package in R^(25)^ (hg38) to determine information related to TE class and family. All analyses were conducted in *limma*^(13–15)^ to evaluate the impact of ATRX loss on gene expression. We adjusted for age, sex, and histology subtype (when relevant) in all models. For analyses of LMS/uLMS, we adjusted for anatomic site (i.e., non-uterine vs. uterine). The empirical Bayes moderated t-statistics were extracted using the *eBayes* function in *limma*. Genes were considered differentially expressed at a nominal p-value < 0.05. We evaluated the distribution TE family and class between *ATRX*-associated TEs and non-*ATRX* associated TEs using FET.

#### Telomere length

Using the previously generated data by Barthel et al. related to telomere length (TL)^(26)^ for TCGA tumors, we examined the impact of ATRX loss on TL dynamics. The tumor and normal TL were estimated from whole genome sequencing or exome sequencing data using the TelSeq software^(27)^, and the ratio of tumor TL/normal TL was utilized in all analyses and log-transformed. Linear models were used to examine impact of *ATRX* on log-transformed tumor TL/normal TL and adjusted for age, sex, histology subtype, and library type (genome vs. exome). We also extracted telomerase signature score provided in the previous study^(26)^ and evaluated using linear models adjusted for age, sex, and histology subtype. All analyses were stratified by histologic subtype. Dichotomized TERRA expression (positive, negative), derived from RNA-seq BAM files using TelSeq^(26)^, was also extracted, and FET was applied to evaluate the association between TERRA expression and ATRX status across all subtypes and within subtypes.

#### Immune infiltration

We obtained immune infiltration cell estimates generated for the PanImmune Atlas for all TCGA samples ^(28)^. These immune cell estimates were generated using Cibersort^(29)^ and expressed as proportion of immune cells. Among the 22 immune cell types available, we analyzed those that were detectable in at least 10% of sarcoma samples. We compared the distribution of the cell type by ATRX status using Mann-U tests. We conducted stratified analyses by sarcoma subtype.

#### Survival analysis

We examined both OS and DFS. We estimated Kaplan-Meier (KM) curves for each of these outcomes and their ATRX loss and extracted the median survival and 5-year survival. Log-rank tests were applied to examine the difference in survival between these curves. To examine the impact of adjusting for other factors related to survival and tumor characteristics, we examined the relationship between OS or DFS using Cox-proportional hazard modeling. Our first set of adjustments include age at diagnosis, sex, and subtype (when relevant), and a second set of adjustments included categorized tumor size (≥ 10 cm, 5-10 cm, and < 5 cm). For analyses of LMS/uLMS, we adjusted for cancer site (non-uterine vs. uterine). The corresponding hazard ratios (HR) and 95% CI were extracted from these models, and p-values were derived from the z-tests. These analyses were stratified by subtype: DDLPS, LMS/uLMS, and UPS/MFS.

All analyses were conducted in R 4.3.3.

### Tables

**Table S1.** ATRX loss of function variants identified among TCGA sarcomas included in analysis (n=49).

| TCGA Patient ID | Sarcoma Histology | Protein Change | Mutation Type |
| --- | --- | --- | --- |
| TCGA-3B-A9HI | Dedifferentiated Liposarcoma (DDLPS) | M828* | ATRX truncating |
| TCGA-3B-A9HO | Dedifferentiated Liposarcoma (DDLPS) |  | Copy number homozygous deletion |
| TCGA-3B-A9HS | Leiomyosarcoma (LMS) | S1253* | ATRX truncating |
| TCGA-3B-A9I1 | Leiomyosarcoma (LMS) | E723* | ATRX truncating |
| TCGA-DX-A1L1 | Dedifferentiated Liposarcoma (DDLPS) |  | Copy number homozygous deletion |
| TCGA-DX-A1L2 | Dedifferentiated Liposarcoma (DDLPS) | V277Gfs*5 | ATRX truncating |
| TCGA-DX-A23R | Dedifferentiated Liposarcoma (DDLPS) |  | Copy number homozygous deletion |
| TCGA-DX-A240 | Dedifferentiated Liposarcoma (DDLPS) |  | Copy number homozygous deletion |
| TCGA-DX-A3LW | Dedifferentiated Liposarcoma (DDLPS) |  | Copy number homozygous deletion |
| TCGA-DX-A3LY | Dedifferentiated Liposarcoma (DDLPS) |  | Copy number homozygous deletion |
| TCGA-DX-A3M2 | Dedifferentiated Liposarcoma (DDLPS) | R1739* | ATRX truncating |
| TCGA-DX-A3U5 | Dedifferentiated Liposarcoma (DDLPS) |  | Copy number homozygous deletion |
| TCGA-DX-A3UD | Leiomyosarcoma (LMS) | R444* | ATRX truncating |
| TCGA-DX-A6B8 | Leiomyosarcoma (LMS) | E1113* | ATRX truncating |
| TCGA-DX-A6YS | Myxofibrosarcoma (MFS) |  | Copy number homozygous deletion |
| TCGA-DX-A7EU | Myxofibrosarcoma (MFS) | L1854Ffs*7 | ATRX truncating |
| TCGA-DX-A8BK | Undifferentiated Pleomorphic Sarcoma (UPS) |  | Structural variant |
| TCGA-DX-A8BM | Undifferentiated Pleomorphic Sarcoma (UPS) | K796Qfs*4 | ATRX truncating |
| TCGA-DX-A8BN | Undifferentiated Pleomorphic Sarcoma (UPS) | Q391* | ATRX truncating |
| TCGA-DX-A8BS | Undifferentiated Pleomorphic Sarcoma (UPS) |  | Copy number homozygous deletion |
| TCGA-DX-A8BT | Undifferentiated Pleomorphic Sarcoma (UPS) | X1439 splice | ATRX truncating |
| TCGA-DX-A8BU | Undifferentiated Pleomorphic Sarcoma (UPS) | N1232Sfs*15 | ATRX truncating |
| TCGA-DX-A8BV | Myxofibrosarcoma (MFS) | K993Nfs*9 | ATRX truncating |
| TCGA-DX-AATS | Undifferentiated Pleomorphic Sarcoma (UPS) | R444Lfs*19 | ATRX truncating |
| TCGA-DX-AB2P | Undifferentiated Pleomorphic Sarcoma (UPS) | M118Yfs*19 | ATRX truncating |
| TCGA-DX-AB2W | Undifferentiated Pleomorphic Sarcoma (UPS) | N2125Wfs*2 | ATRX truncating |
| TCGA-DX-AB37 | Dedifferentiated Liposarcoma (DDLPS) | X1816 splice | ATRX truncating |
| TCGA-DX-AB3A | Dedifferentiated Liposarcoma (DDLPS) | E1024* | ATRX truncating |
| TCGA-FX-A3NK | Dedifferentiated Liposarcoma (DDLPS) |  | Copy number homozygous deletion |
| TCGA-HB-A2OT | Undifferentiated Pleomorphic Sarcoma (UPS) | W263* | ATRX truncating |
| TCGA-HB-A3L4 | Leiomyosarcoma (LMS) | P1750Gfs*4 | ATRX truncating |
| TCGA-HB-A3YV | Undifferentiated Pleomorphic Sarcoma (UPS) |  | Structural variant |
| TCGA-IE-A4EH | Leiomyosarcoma (LMS) | K1409Qfs*80 | ATRX truncating |
| TCGA-IE-A4EJ | Undifferentiated Pleomorphic Sarcoma (UPS) | Q1416*, S1880C | ATRX truncating, ATRX S1880 missense |
| TCGA-JV-A75J | Leiomyosarcoma (LMS) |  | Copy number homozygous deletion |
| TCGA-K1-A6RU | Myxofibrosarcoma (MFS) | S2142Mfs*24 | ATRX truncating |
| TCGA-K1-A6RV | Leiomyosarcoma (LMS) |  | Copy number homozygous deletion |
| TCGA-KD-A5QU | Leiomyosarcoma (LMS) | S1387* | ATRX truncating |
| TCGA-N1-A6IA | Leiomyosarcoma (LMS) |  | Copy number homozygous deletion |
| TCGA-QC-AA9N | Undifferentiated Pleomorphic Sarcoma (UPS) | K806Rfs*16 | ATRX truncating |
| TCGA-SI-AA8B | Undifferentiated Pleomorphic Sarcoma (UPS) | K688* | ATRX truncating |
| TCGA-VT-A80G | Undifferentiated Pleomorphic Sarcoma (UPS) |  | Structural variant |
| TCGA-VT-AB3D | Undifferentiated Pleomorphic Sarcoma (UPS) | E351* | ATRX truncating |
| TCGA-WK-A8Y0 | Undifferentiated Pleomorphic Sarcoma (UPS) | K1701* | ATRX truncating |
| TCGA-WP-A9GB | Leiomyosarcoma (LMS) | E820Ifs*10 | ATRX truncating |
| TCGA-X2-A95T | Leiomyosarcoma (LMS) | K425Rfs*8 | ATRX truncating |
| TCGA-X6-A7WB | Leiomyosarcoma (LMS) |  | Copy number homozygous deletion |
| TCGA-X6-A7WC | Leiomyosarcoma (LMS) | D1618Gfs*8 | ATRX truncating |
| TCGA-X6-A8C6 | Myxofibrosarcoma (MFS) |  | Copy number homozygous deletion |

**Table S2.** Summary of associations with ATRX loss for all analyses and stratified by histologic subtype.

|  | Gene Expression<br>(FDR < 0.05) | Hallmarks GSEA<br>(FDR < 0.05) | Methylation<br>(FDR < 0.05) | TE expression<br>(p < 0.05) | miRNA expression<br>(p < 0.05) |
| --- | --- | --- | --- | --- | --- |
| **All subtypes** | 27 | 11 | 269 | 96 | 37 |
| **DDLPS** | 0 | 17 | 0 | 22 | 5 |
| **LMS/uLMS** | 94 | 26 | 2243 | 134 | 27 |
| **UPS/MFS** | 17 | 11 | 0 | 176 | 29 |

Threshold for associations were either at nominal p-value= 0.05 or false discovery rate (FDR)=0.05. Details regarding each analysis are provided in the *Methods*.

**Table S3.** Differentially expressed genes in sarcomas with ATRX loss (vs. ATRX retained) (FDR < 0.05).

| Ensembl ID | Gene Name | Mean TPM<br>(log<sub>2</sub>-transformed) | logFC (95% CI) | p-value | FDR |
| --- | --- | --- | --- | --- | --- |
| ENSG00000085224.23 | ATRX | 5.70 | -1.08 (-1.41, -0.75) | 9.17E-10 | 1.87E-05 |
| ENSG00000232415.1 | ELN-AS1 | -2.35 | -1.31 (-1.84, -0.77) | 2.36E-06 | 1.62E-02 |
| ENSG00000108591.10 | DRG2 | 4.47 | -0.72 (-1.01, -0.42) | 2.96E-06 | 1.62E-02 |
| ENSG00000280287.1 | AC131212.3 | 0.78 | 0.84 (0.49, 1.18) | 3.17E-06 | 1.62E-02 |
| ENSG00000236264.5 | RPL26P30 | -1.70 | -0.77 (-1.09, -0.44) | 5.23E-06 | 1.86E-02 |
| ENSG00000108679.13 | LGALS3BP | 8.76 | -0.97 (-1.38, -0.56) | 5.90E-06 | 1.86E-02 |
| ENSG00000112941.14 | TENT4A | 5.40 | 0.40 (0.23, 0.56) | 6.39E-06 | 1.86E-02 |
| ENSG00000166246.14 | C16orf71 | 0.17 | -0.83 (-1.19, -0.47) | 1.02E-05 | 2.42E-02 |
| ENSG00000244509.4 | APOBEC3C | 5.68 | -0.67 (-0.97, -0.38) | 1.07E-05 | 2.42E-02 |
| ENSG00000284693.1 | LINC02606 | 0.94 | -0.60 (-0.87, -0.34) | 1.30E-05 | 2.45E-02 |
| ENSG00000245322.7 | H2AZ1-DT | -0.90 | 0.80 (0.45, 1.15) | 1.32E-05 | 2.45E-02 |
| ENSG00000133874.2 | RNF122 | 3.96 | 0.66 (0.37, 0.96) | 1.54E-05 | 2.46E-02 |
| ENSG00000162650.16 | ATXN7L2 | 2.71 | 0.53 (0.29, 0.76) | 1.63E-05 | 2.46E-02 |
| ENSG00000069943.10 | PIGB | 2.17 | -0.48 (-0.70, -0.27) | 1.68E-05 | 2.46E-02 |
| ENSG00000144535.20 | DIS3L2 | 4.27 | -0.37 (-0.54, -0.20) | 2.07E-05 | 2.83E-02 |
| ENSG00000196632.10 | WNK3 | 1.19 | 1.12 (0.61, 1.63) | 2.48E-05 | 3.14E-02 |
| ENSG00000116096.6 | SPR | 4.06 | -0.74 (-1.08, -0.40) | 2.61E-05 | 3.14E-02 |
| ENSG00000130595.19 | TNNT3 | 1.81 | 1.81 (0.96, 2.66) | 3.75E-05 | 4.17E-02 |
| ENSG00000286366.1 | AC005052.2 | -1.04 | -0.59 (-0.87, -0.31) | 4.34E-05 | 4.17E-02 |
| ENSG00000162642.14 | C1orf52 | 4.38 | 0.34 (0.18, 0.50) | 4.40E-05 | 4.17E-02 |
| ENSG00000132622.11 | HSPA12B | 4.65 | 0.90 (0.47, 1.32) | 4.64E-05 | 4.17E-02 |
| ENSG00000184185.10 | KCNJ12 | 2.42 | 1.28 (0.67, 1.89) | 4.67E-05 | 4.17E-02 |
| ENSG00000123595.8 | RAB9A | 4.83 | 0.42 (0.22, 0.63) | 4.69E-05 | 4.17E-02 |
| ENSG00000204764.14 | RANBP17 | 1.10 | 1.22 (0.64, 1.81) | 5.31E-05 | 4.52E-02 |
| ENSG00000142530.11 | FAM71E1 | -1.44 | -1.13 (-1.67, -0.58) | 6.35E-05 | 4.99E-02 |
| ENSG00000237807.4 | AC022034.1 | 1.26 | -1.89 (-2.81, -0.98) | 6.45E-05 | 4.99E-02 |
| ENSG00000112139.16 | MDGA1 | 4.03 | 0.82 (0.42, 1.22) | 6.59E-05 | 4.99E-02 |

Results obtained from linear regression models conducted in limma (see *Methods* for more details). Models adjusted for age at diagnosis, sex, and histologic subtype. P-values obtained from moderated t-statistic.

**Table S4.** Differentially expressed genes in Undifferentiated Pleomorphic Sarcoma (UPS) and Myxofibrosarcoma (MFS) with ATRX loss (vs. ATRX retained) (FDR < 0.05).

| Ensembl ID | Gene Name | Mean TPM<br>(log<sub>2</sub>-transformed) | logFC (95% CI) | p-value | FDR |
| --- | --- | --- | --- | --- | --- |
| ENSG00000085224.23 | ATRX | 5.48 | -1.21 (-1.63, -0.79) | 2.24E-07 | 0.0046 |
| ENSG00000285210.1 | LINC02795 | -2.09 | 1.59 ( 0.96, 2.22) | 2.87E-06 | 0.022 |
| ENSG00000196632.10 | WNK3 | 0.74 | 2.09 ( 1.26, 2.91) | 3.18E-06 | 0.022 |
| ENSG00000145248.7 | SLC10A4 | -0.89 | 2.42 ( 1.42, 3.41) | 6.85E-06 | 0.028 |
| ENSG00000108679.13 | LGALS3BP | 8.98 | -1.36 (-1.92, -0.80) | 7.05E-06 | 0.028 |
| ENSG00000172426.16 | RSPH9 | -0.17 | 1.46 ( 0.86, 2.07) | 8.16E-06 | 0.028 |
| ENSG00000135097.7 | MSI1 | -1.43 | 3.07 ( 1.76, 4.37) | 1.27E-05 | 0.037 |
| ENSG00000003989.18 | SLC7A2 | 3.30 | 1.71 ( 0.97, 2.46) | 1.85E-05 | 0.041 |
| ENSG00000214694.12 | ARHGEF33 | -0.86 | 1.44 ( 0.81, 2.07) | 1.91E-05 | 0.041 |
| ENSG00000182771.19 | GRID1 | 1.44 | 2.22 ( 1.24, 3.19) | 2.17E-05 | 0.041 |
| ENSG00000133874.2 | RNF122 | 3.82 | 0.96 ( 0.53, 1.39) | 2.75E-05 | 0.041 |
| ENSG00000184402.15 | SS18L1 | 4.15 | 0.68 ( 0.38, 0.98) | 2.99E-05 | 0.041 |
| ENSG00000185619.18 | PCGF3 | 5.14 | 0.68 ( 0.38, 0.99) | 3.04E-05 | 0.041 |
| ENSG00000132639.13 | SNAP25 | -0.12 | 2.45 ( 1.35, 3.55) | 3.15E-05 | 0.041 |
| ENSG00000204950.4 | LRRC10B | 0.14 | 1.89 ( 1.04, 2.75) | 3.18E-05 | 0.041 |
| ENSG00000101605.13 | MYOM1 | 2.25 | 1.42 ( 0.78, 2.06) | 3.24E-05 | 0.041 |
| ENSG00000188517.16 | COL25A1 | -0.56 | 2.39 ( 1.31, 3.47) | 3.64E-05 | 0.044 |

Results obtained from linear regression models conducted in limma (see *Methods* for more details). Models adjusted for age at diagnosis, sex, and histologic subtype (e.g., MFS, UPS). P-values obtained from moderated t-statistic.

**Table S5.**
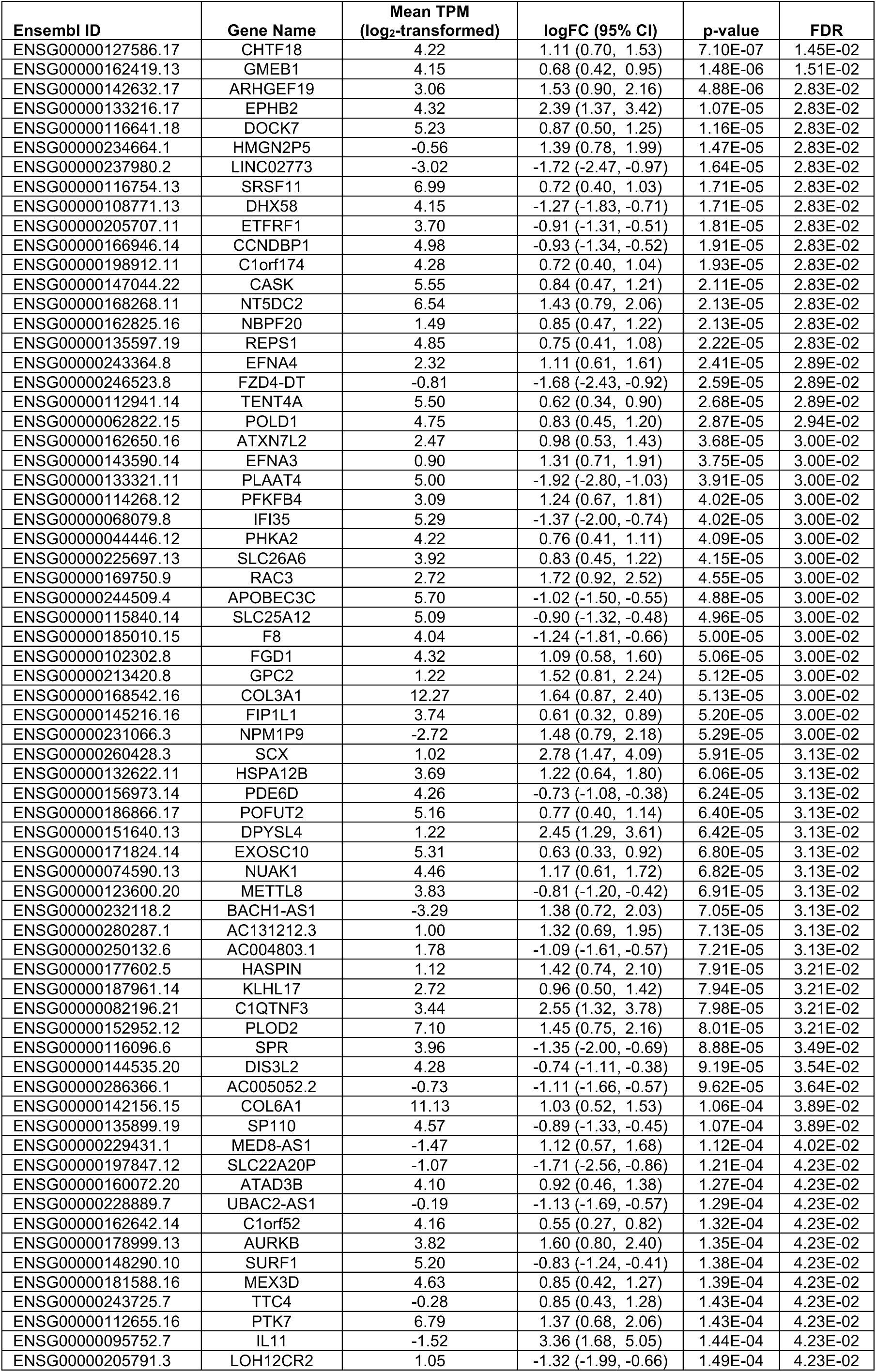
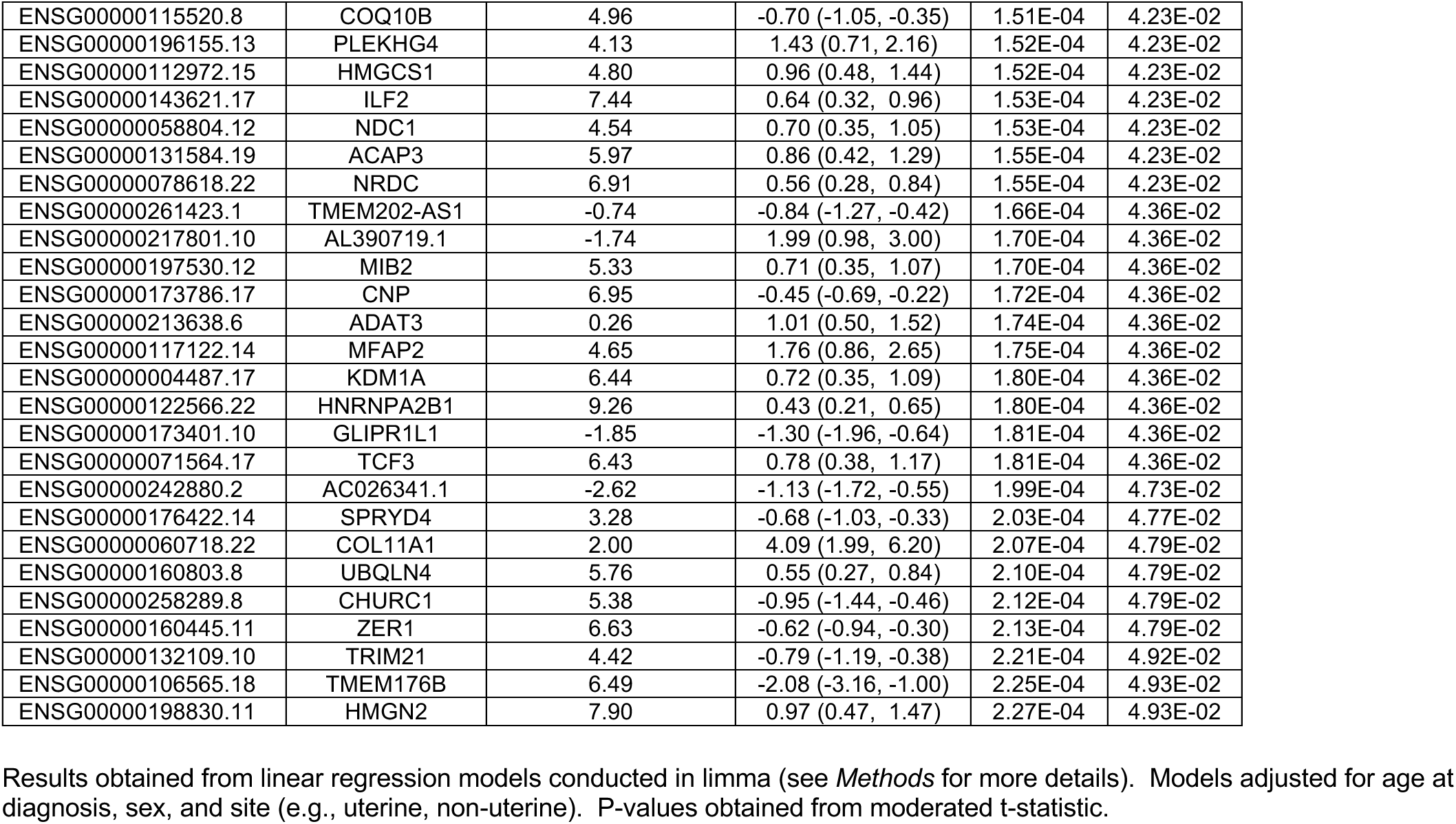
Differentially expressed genes in Leiomyosarcoma (LMS) with ATRX loss (vs. ATRX retained) (FDR < 0.05).

**Table S6.** Impact of ATRX loss on normalized enrichment scores (NES) for Hallmark gene sets.

| Hallmark Term | Set Size | All Sarcomas |  |  |  | DDLPS |  |  |  | LMS/uLMS |  |  |  | UPS/MFS |  |  |  |
| --- | --- | --- | --- | --- | --- | --- | --- | --- | --- | --- | --- | --- | --- | --- | --- | --- | --- |
|  |  | NES | p | FDR | % | NES | p | FDR | % | NES | p | FDR | % | NES | p | FDR | % |
| ADIPOGENESIS | 190 | -1.04 | 3.7E-01 | 6.2E-01 | 34% | 1.30 | 3.0E-02 | 7.8E-02 | 22% | -1.65 | 1.5E-04 | **5.3E-04** | 35% | 0.62 | 1.0E+00 | 1.0E+00 | 11% |
| ALLOGRAFT REJECTION | 185 | -2.02 | 1.0E-08 | **1.0E-07** | 39% | 1.98 | 8.4E-08 | **1.4E-06** | 43% | -1.99 | 1.5E-08 | **9.6E-08** | 42% | -2.52 | 7.4E-16 | **1.2E-14** | 46% |
| ANDROGEN RESPONSE | 96 | -1.03 | 3.8E-01 | 6.2E-01 | 17% | -0.69 | 9.7E-01 | 1.0E+00 | 20% | 0.93 | 6.1E-01 | 6.5E-01 | 25% | -1.17 | 1.5E-01 | 2.9E-01 | 25% |
| ANGIOGENESIS | 31 | 1.28 | 1.5E-01 | 3.9E-01 | 42% | 1.16 | 2.3E-01 | 3.8E-01 | 16% | 1.27 | 1.2E-01 | 2.0E-01 | 26% | 1.47 | 4.5E-02 | 1.2E-01 | 48% |
| APICAL JUNCTION | 184 | -1.14 | 1.7E-01 | 4.1E-01 | 17% | -1.37 | 1.8E-02 | 5.3E-02 | 20% | 1.27 | 4.5E-02 | 8.0E-02 | 22% | 1.26 | 7.2E-02 | 1.6E-01 | 29% |
| APICAL SURFACE | 39 | -1.03 | 4.2E-01 | 6.2E-01 | 18% | -1.26 | 1.5E-01 | 2.9E-01 | 28% | 1.15 | 2.1E-01 | 3.1E-01 | 31% | 1.02 | 4.3E-01 | 5.9E-01 | 26% |
| APOPTOSIS | 154 | 1.04 | 3.3E-01 | 6.2E-01 | 23% | 1.53 | 2.5E-03 | **9.8E-03** | 29% | -0.82 | 8.7E-01 | 8.9E-01 | 19% | -1.16 | 1.7E-01 | 3.1E-01 | 24% |
| BILE ACID METABOLISM | 96 | -1.04 | 3.7E-01 | 6.2E-01 | 25% | 1.27 | 8.0E-02 | 1.7E-01 | 29% | -1.88 | 4.1E-05 | **1.7E-04** | 40% | 1.09 | 3.0E-01 | 4.7E-01 | 25% |
| CHOLESTEROL HOMEOSTASIS | 71 | 1.04 | 3.8E-01 | 6.2E-01 | 32% | 0.72 | 9.6E-01 | 1.0E+00 | 30% | 1.40 | 1.7E-02 | **3.3E-02** | 46% | -1.00 | 4.4E-01 | 5.9E-01 | 21% |
| COAGULATION | 106 | -0.93 | 6.0E-01 | 8.1E-01 | 23% | 1.62 | 3.1E-03 | **1.1E-02** | 36% | -1.10 | 3.0E-01 | 4.1E-01 | 22% | -1.02 | 4.3E-01 | 5.9E-01 | 26% |
| COMPLEMENT | 184 | -1.30 | 3.3E-02 | 1.2E-01 | 26% | 1.80 | 8.9E-06 | **1.1E-04** | 40% | -1.52 | 2.4E-03 | **6.2E-03** | 28% | -1.72 | 4.6E-05 | **2.9E-04** | 24% |
| DNA REPAIR | 148 | -0.44 | 1.0E+00 | 1.0E+00 | 23% | 1.01 | 4.1E-01 | 5.9E-01 | 51% | 1.02 | 3.7E-01 | 4.8E-01 | 35% | -1.37 | 1.6E-02 | 6.5E-02 | 39% |
| E2F TARGETS | 199 | 2.29 | 2.5E-13 | **4.2E-12** | 45% | 1.15 | 1.4E-01 | 2.9E-01 | 33% | 2.92 | 1.1E-28 | **5.5E-27** | 62% | -1.12 | 1.8E-01 | 3.2E-01 | 39% |
| EPITHELIAL MESENCHYMAL TRANSITION | 197 | 1.86 | 3.8E-07 | **2.7E-06** | 34% | -1.54 | 2.0E-03 | **9.8E-03** | 30% | 2.61 | 3.0E-19 | **5.0E-18** | 44% | 1.33 | 2.5E-02 | 8.8E-02 | 35% |
| ESTROGEN RESPONSE EARLY | 190 | -1.58 | 9.7E-04 | **6.1E-03** | 21% | -1.26 | 6.7E-02 | 1.6E-01 | 31% | -1.41 | 1.1E-02 | **2.2E-02** | 26% | -1.23 | 5.1E-02 | 1.3E-01 | 19% |
| ESTROGEN RESPONSE LATE | 180 | -1.30 | 4.8E-02 | 1.6E-01 | 27% | 1.41 | 1.4E-02 | **4.4E-02** | 23% | -1.32 | 4.0E-02 | 7.4E-02 | 30% | -1.22 | 6.4E-02 | 1.5E-01 | 34% |
| FATTY ACID METABOLISM | 142 | 0.76 | 9.7E-01 | 1.0E+00 | 20% | 1.60 | 9.6E-04 | **5.3E-03** | 32% | -1.48 | 6.7E-03 | **1.5E-02** | 37% | 0.72 | 9.6E-01 | 1.0E+00 | 21% |
| G2M CHECKPOINT | 199 | 2.04 | 2.6E-09 | **3.3E-08** | 44% | 0.96 | 5.4E-01 | 7.1E-01 | 32% | 2.78 | 1.3E-23 | **3.2E-22** | 50% | -1.10 | 2.1E-01 | 3.6E-01 | 41% |
| GLYCOLYSIS | 186 | 1.41 | 5.4E-03 | **2.4E-02** | 33% | -0.96 | 5.6E-01 | 7.1E-01 | 18% | 1.91 | 1.8E-07 | **9.1E-07** | 30% | 0.98 | 4.9E-01 | 6.5E-01 | 18% |
| HEDGEHOG SIGNALING | 34 | 1.04 | 4.0E-01 | 6.2E-01 | 41% | -1.79 | 2.4E-03 | **9.8E-03** | 38% | 1.18 | 2.0E-01 | 3.1E-01 | 18% | 1.66 | 1.1E-02 | 5.3E-02 | 47% |
| HEME METABOLISM | 178 | -0.75 | 9.8E-01 | 1.0E+00 | 28% | 0.87 | 8.4E-01 | 1.0E+00 | 16% | -1.11 | 2.4E-01 | 3.4E-01 | 31% | 0.74 | 9.5E-01 | 1.0E+00 | 23% |
| HYPOXIA | 187 | 1.15 | 1.5E-01 | 3.9E-01 | 26% | -1.24 | 9.8E-02 | 2.0E-01 | 24% | 1.45 | 4.5E-03 | **1.1E-02** | 30% | 1.25 | 8.0E-02 | 1.7E-01 | 29% |
| IL2 STAT5 SIGNALING | 194 | -1.22 | 8.5E-02 | 2.6E-01 | 22% | 1.08 | 3.0E-01 | 4.7E-01 | 28% | -1.00 | 4.5E-01 | 5.3E-01 | 30% | -1.37 | 1.2E-02 | 5.5E-02 | 27% |
| IL6 JAK STAT3 SIGNALING | 80 | -1.02 | 4.2E-01 | 6.2E-01 | 29% | 1.75 | 9.6E-04 | **5.3E-03** | 36% | -1.61 | 2.9E-03 | **7.2E-03** | 40% | -1.34 | 4.5E-02 | 1.2E-01 | 19% |
| INFLAMMATORY RESPONSE | 183 | -1.54 | 1.9E-03 | **1.0E-02** | 29% | 1.78 | 1.3E-05 | **1.3E-04** | 39% | -1.45 | 6.8E-03 | **1.5E-02** | 31% | -1.99 | 1.7E-07 | **1.7E-06** | 33% |
| INTERFERON ALPHA RESPONSE | 97 | -2.51 | 3.4E-14 | **8.4E-13** | 69% | 2.44 | 1.1E-10 | **2.7E-09** | 58% | -2.38 | 1.4E-11 | **1.4E-10** | 70% | -3.06 | 6.3E-24 | **1.6E-22** | 70% |
| INTERFERON GAMMA RESPONSE | 196 | -2.48 | 1.3E-18 | **6.4E-17** | 51% | 2.29 | 9.6E-13 | **4.8E-11** | 52% | -2.36 | 4.4E-16 | **5.5E-15** | 61% | -3.10 | 5.8E-34 | **2.9E-32** | 57% |
| KRAS SIGNALING DN | 122 | -1.43 | 1.2E-02 | **4.9E-02** | 30% | -1.73 | 5.8E-04 | **4.1E-03** | 28% | -1.59 | 2.1E-03 | **6.1E-03** | 30% | 1.54 | 3.6E-03 | **2.0E-02** | 35% |
| KRAS SIGNALING UP | 178 | -1.10 | 2.5E-01 | 5.6E-01 | 19% | 1.15 | 1.7E-01 | 3.1E-01 | 26% | -1.02 | 4.3E-01 | 5.1E-01 | 24% | -1.08 | 2.4E-01 | 4.0E-01 | 14% |
| MITOTIC SPINDLE | 198 | 1.89 | 1.5E-07 | **1.2E-06** | 49% | 0.61 | 1.0E+00 | 1.0E+00 | 28% | 2.15 | 2.4E-10 | **2.0E-09** | 35% | 0.94 | 6.1E-01 | 7.7E-01 | 44% |
| MTORC1 SIGNALING | 199 | 0.89 | 7.7E-01 | 9.6E-01 | 23% | -0.84 | 8.4E-01 | 1.0E+00 | 22% | 1.72 | 1.2E-05 | **5.3E-05** | 31% | -1.27 | 3.1E-02 | 9.7E-02 | 35% |
| MYC TARGETS V1 | 199 | 0.74 | 9.9E-01 | 1.0E+00 | 27% | -0.49 | 1.0E+00 | 1.0E+00 | 31% | 2.01 | 9.4E-09 | **6.7E-08** | 54% | -1.86 | 5.1E-07 | **4.2E-06** | 55% |
| MYC TARGETS V2 | 57 | -0.85 | 7.5E-01 | 9.6E-01 | 53% | 0.72 | 9.5E-01 | 1.0E+00 | 30% | 1.67 | 2.1E-03 | **6.1E-03** | 51% | -2.22 | 1.1E-06 | **8.1E-06** | 60% |
| MYOGENESIS | 175 | -1.37 | 1.7E-02 | 6.6E-02 | 23% | -1.01 | 4.1E-01 | 5.9E-01 | 30% | -1.97 | 1.3E-07 | **7.5E-07** | 37% | 1.92 | 1.3E-07 | **1.7E-06** | 40% |
| NOTCH SIGNALING | 32 | 0.67 | 9.2E-01 | 1.0E+00 | 47% | -0.70 | 9.0E-01 | 1.0E+00 | 41% | 0.87 | 7.0E-01 | 7.2E-01 | 34% | 0.81 | 7.6E-01 | 9.3E-01 | 34% |
| OXIDATIVE PHOSPHORYLATION | 200 | -1.44 | 4.1E-03 | **2.0E-02** | 48% | 1.30 | 2.9E-02 | 7.8E-02 | 48% | -1.68 | 1.4E-04 | **5.3E-04** | 57% | -1.29 | 2.6E-02 | 8.8E-02 | 46% |
| P53 PATHWAY | 191 | 0.78 | 9.8E-01 | 1.0E+00 | 34% | 1.13 | 2.0E-01 | 3.4E-01 | 24% | 0.97 | 5.8E-01 | 6.5E-01 | 26% | -0.96 | 6.1E-01 | 7.7E-01 | 33% |
| PANCREAS BETA CELLS | 19 | 1.34 | 1.2E-01 | 3.6E-01 | 26% | -1.46 | 7.0E-02 | 1.6E-01 | 21% | 1.21 | 2.1E-01 | 3.1E-01 | 21% | 1.34 | 1.1E-01 | 2.2E-01 | 21% |
| PEROXISOME | 94 | 0.76 | 9.5E-01 | 1.0E+00 | 19% | 1.03 | 3.8E-01 | 5.8E-01 | 28% | -1.06 | 3.4E-01 | 4.5E-01 | 32% | 0.72 | 9.4E-01 | 1.0E+00 | 26% |
| PI3K AKT MTOR SIGNALING | 101 | -0.88 | 6.8E-01 | 9.0E-01 | 9% | 1.06 | 3.0E-01 | 4.7E-01 | 30% | -0.92 | 6.1E-01 | 6.5E-01 | 10% | -0.58 | 1.0E+00 | 1.0E+00 | 38% |
| PROTEIN SECRETION | 94 | -0.60 | 1.0E+00 | 1.0E+00 | 5% | -0.99 | 4.7E-01 | 6.4E-01 | 29% | -0.54 | 1.0E+00 | 1.0E+00 | 9% | 0.67 | 9.7E-01 | 1.0E+00 | 37% |
| REACTIVE OXYGEN SPECIES PATHWAY | 48 | 0.74 | 9.0E-01 | 1.0E+00 | 17% | 1.60 | 1.4E-02 | **4.4E-02** | 46% | -1.10 | 3.4E-01 | 4.5E-01 | 38% | 0.65 | 9.7E-01 | 1.0E+00 | 8% |
| SPERMATOGENESIS | 94 | 1.16 | 1.6E-01 | 3.9E-01 | 18% | -0.62 | 1.0E+00 | 1.0E+00 | 31% | 0.94 | 6.1E-01 | 6.5E-01 | 22% | 1.38 | 3.7E-02 | 1.1E-01 | 22% |
| TGF BETA SIGNALING | 52 | 1.09 | 2.8E-01 | 5.8E-01 | 25% | -0.57 | 9.9E-01 | 1.0E+00 | 37% | 1.19 | 1.8E-01 | 2.8E-01 | 27% | 0.76 | 8.5E-01 | 1.0E+00 | 17% |
| TNFA SIGNALING VIA NFKB | 193 | 1.01 | 4.1E-01 | 6.2E-01 | 23% | 1.30 | 4.0E-02 | 1.0E-01 | 29% | 1.18 | 9.2E-02 | 1.6E-01 | 19% | -1.15 | 9.9E-02 | 2.1E-01 | 19% |
| UNFOLDED PROTEIN RESPONSE | 111 | 0.95 | 5.5E-01 | 7.9E-01 | 34% | -0.68 | 9.9E-01 | 1.0E+00 | 32% | 1.60 | 2.0E-03 | **6.1E-03** | 48% | -0.66 | 1.0E+00 | 1.0E+00 | 38% |
| UV RESPONSE DN | 144 | 0.95 | 5.8E-01 | 8.0E-01 | 24% | -1.54 | 2.4E-03 | **9.8E-03** | 24% | 1.36 | 1.5E-02 | **3.1E-02** | 20% | 1.03 | 4.2E-01 | 5.9E-01 | 24% |
| UV RESPONSE UP | 152 | 1.05 | 3.2E-01 | 6.2E-01 | 20% | 1.01 | 4.4E-01 | 6.1E-01 | 22% | -1.03 | 4.0E-01 | 5.1E-01 | 26% | 1.39 | 2.3E-02 | 8.8E-02 | 20% |
| WNT BETA CATENIN SIGNALING | 40 | 0.70 | 9.1E-01 | 1.0E+00 | 25% | -1.23 | 1.7E-01 | 3.1E-01 | 38% | 1.02 | 4.1E-01 | 5.1E-01 | 28% | 1.13 | 3.0E-01 | 4.7E-01 | 35% |
| XENOBIOTIC METABOLISM | 168 | -1.08 | 2.7E-01 | 5.8E-01 | 17% | 1.69 | 1.0E-04 | **8.7E-04** | 31% | -1.57 | 2.2E-03 | **6.1E-03** | 35% | 1.07 | 3.4E-01 | 5.1E-01 | 18% |

**Table S7.** Impact of ATRX loss on estimates of tumor immune infiltrates from Cibersort.

|  | All Sarcomas |  |  | DDLPS |  |  | LMS (all sites) |  |  | UPS/MFS |  |  |
| --- | --- | --- | --- | --- | --- | --- | --- | --- | --- | --- | --- | --- |
| Immune Infiltrate | ATRX retained | ATRX loss | p | ATRX retained | ATRX loss | p | ATRX retained | ATRX loss | p | ATRX retained | ATRX loss | p |
| **B-cells** | 0.02 (0.01, 0.05) | 0.01 (0.01, 0.03) | 0.2490 | 0.02 (0.01, 0.04) | 0.02 (0.01, 0.07) | 0.6146 | 0.03 (0.01, 0.06) | 0.01 (0.00, 0.02) | **0.0065** | 0.02 (0.00, 0.03) | 0.02 (0.01, 0.03) | 0.3988 |
| B.cells.memory | 0.00 (0.00, 0.01) | 0.00 (0.00, 0.00) | 0.0741 | 0.00 (0.00, 0.00) | 0.00 (0.00, 0.00) | 0.2658 | 0.00 (0.00, 0.02) | 0.00 (0.00, 0.01) | 0.5433 | 0.00 (0.00, 0.01) | 0.00 (0.00, 0.01) | 0.2939 |
| B.cells.naive | 0.00 (0.00, 0.02) | 0.01 (0.00, 0.02) | 0.7069 | 0.01 (0.00, 0.03) | 0.01 (0.00, 0.06) | 0.7360 | 0.00 (0.00, 0.02) | 0.00 (0.00, 0.01) | 0.1061 | 0.00 (0.00, 0.01) | 0.01 (0.00, 0.02) | **0.0364** |
| Plasma.cells | 0.00 (0.00, 0.02) | 0.00 (0.00, 0.01) | 0.6931 | 0.00 (0.00, 0.01) | 0.00 (0.00, 0.02) | 0.3029 | 0.00 (0.00, 0.02) | 0.00 (0.00, 0.00) | 0.0766 | 0.00 (0.00, 0.01) | 0.00 (0.00, 0.01) | 0.1356 |
| **CD4-T cells** | 0.13 (0.09, 0.18) | 0.09 (0.06, 0.17) | **0.0322** | 0.13 (0.11, 0.17) | 0.08 (0.06, 0.12) | **0.0074** | 0.16 (0.11, 0.20) | 0.18 (0.12, 0.20) | 0.6695 | 0.10 (0.06, 0.13) | 0.07 (0.04, 0.15) | 0.5646 |
| T.cells.CD4.memory.activated | 0.00 (0.00, 0.00) | 0.00 (0.00, 0.00) | 0.3933 | 0.00 (0.00, 0.00) | 0.00 (0.00, 0.00) | 0.5137 | 0.00 (0.00, 0.00) | 0.00 (0.00, 0.00) | 0.8440 | 0.00 (0.00, 0.00) | 0.00 (0.00, 0.00) | 0.2201 |
| T.cells.CD4.memory.resting | 0.09 (0.05, 0.14) | 0.06 (0.02, 0.13) | **0.0460** | 0.12 (0.08, 0.15) | 0.05 (0.03, 0.08) | **0.0033** | 0.12 (0.05, 0.17) | 0.15 (0.07, 0.18) | 0.5771 | 0.05 (0.01, 0.08) | 0.05 (0.01, 0.08) | 0.9535 |
| T.cells.CD4.naive | 0.00 (0.00, 0.00) | 0.00 (0.00, 0.00) | 0.9714 | 0.00 (0.00, 0.00) | 0.00 (0.00, 0.00) | 0.4610 | 0.00 (0.00, 0.00) | 0.00 (0.00, 0.00) | 0.3613 | 0.00 (0.00, 0.00) | 0.00 (0.00, 0.00) | **0.0286** |
| T.cells.follicular.helper | 0.02 (0.01, 0.05) | 0.03 (0.01, 0.04) | 0.3091 | 0.01 (0.00, 0.04) | 0.03 (0.01, 0.05) | 0.1000 | 0.02 (0.01, 0.05) | 0.03 (0.02, 0.06) | 0.2711 | 0.03 (0.01, 0.05) | 0.02 (0.01, 0.04) | 0.4959 |
| **Macrophage** | 0.50 (0.36, 0.63) | 0.61 (0.46, 0.73) | **0.0028** | 0.55 (0.41, 0.66) | 0.59 (0.48, 0.72) | 0.2749 | 0.42 (0.32, 0.52) | 0.46 (0.38, 0.60) | 0.3635 | 0.58 (0.43, 0.73) | 0.66 (0.56, 0.75) | 0.1021 |
| Macrophages.M0 | 0.00 (0.00, 0.05) | 0.00 (0.00, 0.15) | 0.1167 | 0.00 (0.00, 0.02) | 0.00 (0.00, 0.00) | 0.4946 | 0.00 (0.00, 0.04) | 0.09 (0.03, 0.19) | **0.0015** | 0.00 (0.00, 0.11) | 0.00 (0.00, 0.15) | 0.8845 |
| Macrophages.M1 | 0.03 (0.01, 0.06) | 0.02 (0.00, 0.05) | 0.2761 | 0.03 (0.01, 0.07) | 0.03 (0.02, 0.07) | 0.9257 | 0.02 (0.01, 0.06) | 0.01 (0.00, 0.03) | 0.1226 | 0.03 (0.01, 0.08) | 0.03 (0.00, 0.06) | 0.4179 |
| Macrophages.M2 | 0.39 (0.29, 0.52) | 0.47 (0.34, 0.59) | **0.0181** | 0.47 (0.33, 0.58) | 0.51 (0.41, 0.58) | 0.2832 | 0.35 (0.26, 0.44) | 0.34 (0.31, 0.42) | 0.9960 | 0.47 (0.34, 0.59) | 0.55 (0.43, 0.63) | 0.1260 |
| **Mast** | 0.05 (0.03, 0.14) | 0.06 (0.03, 0.09) | 0.4841 | 0.04 (0.02, 0.08) | 0.05 (0.03, 0.07) | 1.0000 | 0.11 (0.04, 0.20) | 0.08 (0.05, 0.13) | 0.4544 | 0.04 (0.02, 0.06) | 0.06 (0.02, 0.08) | 0.2766 |
| Mast.cells.activated | 0.00 (0.00, 0.00) | 0.00 (0.00, 0.00) | **0.0154** | 0.00 (0.00, 0.00) | 0.00 (0.00, 0.00) | 0.9628 | 0.00 (0.00, 0.00) | 0.00 (0.00, 0.02) | 0.1575 | 0.00 (0.00, 0.00) | 0.00 (0.00, 0.01) | **0.0220** |
| Mast.cells.resting | 0.05 (0.02, 0.14) | 0.05 (0.00, 0.08) | 0.1112 | 0.04 (0.01, 0.08) | 0.05 (0.03, 0.07) | 0.6811 | 0.11 (0.04, 0.19) | 0.06 (0.01, 0.13) | 0.2073 | 0.04 (0.01, 0.06) | 0.05 (0.00, 0.08) | 0.8018 |
| **Monocytes** | 0.04 (0.01, 0.07) | 0.03 (0.01, 0.06) | 0.2522 | 0.04 (0.02, 0.06) | 0.02 (0.01, 0.08) | 0.5757 | 0.04 (0.02, 0.07) | 0.03 (0.00, 0.04) | 0.0922 | 0.02 (0.01, 0.06) | 0.03 (0.01, 0.06) | 0.6444 |
| **NK** | 0.03 (0.02, 0.06) | 0.04 (0.02, 0.06) | 0.9780 | 0.03 (0.01, 0.05) | 0.04 (0.02, 0.05) | 0.2048 | 0.04 (0.02, 0.06) | 0.05 (0.02, 0.07) | 0.5369 | 0.03 (0.02, 0.06) | 0.03 (0.01, 0.04) | 0.3283 |
| NK.cells.activated | 0.02 (0.01, 0.05) | 0.02 (0.01, 0.04) | 0.8467 | 0.02 (0.01, 0.04) | 0.03 (0.02, 0.04) | 0.2107 | 0.03 (0.01, 0.05) | 0.03 (0.01, 0.05) | 0.6619 | 0.02 (0.01, 0.05) | 0.02 (0.00, 0.04) | 0.4138 |
| NK.cells.resting | 0.00 (0.00, 0.01) | 0.00 (0.00, 0.01) | 0.9593 | 0.00 (0.00, 0.00) | 0.00 (0.00, 0.00) | 0.5242 | 0.00 (0.00, 0.01) | 0.00 (0.00, 0.02) | 0.7663 | 0.00 (0.00, 0.01) | 0.00 (0.00, 0.01) | 0.6875 |
| **T.cells.CD8** | 0.07 (0.04, 0.13) | 0.06 (0.02, 0.14) | 0.2109 | 0.06 (0.04, 0.10) | 0.06 (0.03, 0.11) | 0.6583 | 0.07 (0.04, 0.12) | 0.06 (0.02, 0.15) | 0.6622 | 0.08 (0.04, 0.17) | 0.06 (0.02, 0.11) | 0.2790 |
| **T.cells.regulatory..Tregs.** | 0.02 (0.00, 0.04) | 0.01 (0.00, 0.02) | 0.1683 | 0.02 (0.00, 0.03) | 0.01 (0.00, 0.03) | 0.7499 | 0.01 (0.00, 0.03) | 0.02 (0.00, 0.02) | 0.9267 | 0.02 (0.01, 0.04) | 0.01 (0.00, 0.02) | **0.0149** |

Estimates of immune infiltrates obtained from previously generated estimates from Cibersort (see *Methods* for more detail). Medians and interquartile range (25<sup>th</sup>, 75<sup>th</sup> percentile) presented for ATRX retained and loss sarcomas within each histologic subtype grouping (or overall). P-values obtained from Mann-U test. Bolded values indicate p < 0.05.

**Table S8.** Impact of ATRX loss on overall and disease-free survival among sarcoma patients in TCGA.

|  | 5-year Survival (95% CI) |  | Median survival |  |  | Model 1 |  | Model 2 |  |
| --- | --- | --- | --- | --- | --- | --- | --- | --- | --- |
|  | ATRX retained | ATRX Loss | ATRX retained | ATRX Loss | p<sub>log-rank</sub> | HR (95% CI) | p-value | HR (95% CI) | p-value |
| OVERALL SURVIVAL |  |  |  |  |  |  |  |  |  |
| All tumors (n=234) | 56.3 (47.7, 66.5) | 47.5 (33.5, 67.4) | 76.4 | 54.2 | 0.25 | 1.32 (0.80, 2.18) | 0.28 | 1.27 (0.70, 2.29) | 0.43 |
| DDLPS (n=59) | 41.5 (26.7, 64.5) | 75.0 (49.6, 100.0) | 37.1 | 84.7 | 0.028 | 0.29 (0.08, 1.01) | 0.051 | 0.25 (0.06, 1.11) | 0.068 |
| LMS/uLMS (n=100) | 62.4 (50.7, 76.7) | 27.1 (10.5, 69.6) | 81.0 | 34.9 | 0.0023 | 3.63 (1.62, 8.14) | 0.0017 | 2.89 (1.11, 7.49) | 0.030 |
| uLMS (uterine, n=29) | 58.0 (37.8, 89.1) | 14.8 (2.6, 86.0) | 63.8 | 18.6 | 0.11 | 5.34 (1.35, 21.12) | 0.017 | 5.18 (1.06, 25.30) | 0.042 |
| LMS (not uterine, n=71) | 63.0 (49.5, 80.2) | 50.0 (18.8, 100.0) | 81.0 | 44.9 | 0.36 | 2.13 (0.43, 10.53) | 0.35 | 1.12 (0.13, 9.48) | 0.92 |
| UPS/MFS (n=75) | 56.8 (40.5, 79.7) | 50.4 (31.5, 80.6) | NA | 60.7 | 0.29 | 1.86 (0.80, 4.34) | 0.15 | 1.43 (0.48, 4.23) | 0.52 |
| UPS (n=50) | 49.3 (27.6, 87.8) | 49.8 (28.9, 85.9) | 46.8 | 36.7 | 0.46 | 2.10 (0.72, 6.15) | 0.18 | 1.80 (0.43, 7.62) | 0.42 |
| MFS (n=25) | 66.4 (45.4, 97.2) | 53.3 (21.4, 100.0) | NA | 60.7 | 0.41 | 2.58 (0.58, 11.55) | 0.21 | 59.20 (0.92, 3799.26) | 0.055 |
| DISEASE-FREE SURVIVAL |  |  |  |  |  |  |  |  |  |
| All tumors (n=234) | 39.1 (31.5, 48.4) | 34.5 (21.9, 54.4) | 27.2 | 19.9 | 0.76 | 1.03 (0.66, 1.61) | 0.90 | 1.02 (0.62, 1.68) | 0.94 |
| DDLPS (n=59) | 25.2 (13.2, 48.2) | 40.3 (17.8, 91.2) | 18.7 | 48.7 | 0.075 | 0.45 (0.18, 1.10) | 0.078 | 0.31 (0.10, 0.93) | 0.036 |
| LMS all (n=100) | 43.4 (32.9, 57.1) | 12.1 (2.2, 65.7) | 29.4 | 7.0 | 0.011 | 2.30 (1.13, 4.70) | 0.022 | 1.67 (0.73, 3.80) | 0.22 |
| LMS (uterine, n=29) | 29.8 (12.4, 71.8) | 16.7 (3.2, 88.2) | 28.2 | 7.0 | 0.49 | 3.61 (1.05, 12.49) | 0.042 | 3.45 (0.88, 13.60) | 0.077 |
| LMS (not uterine, n=71) | 45.9 (34.2, 61.7) | 0.0 (0.0, 0.0) | 31.7 | 9.9 | 0.012 | 4.24 (1.34, 13.44) | 0.014 | 2.50 (0.64, 9.79) | 0.19 |
| UPS/MFS (n=75) | 43.3 (30.1, 62.5) | 48.7 (30.7, 77.4) | 25.8 | 22.0 | 0.90 | 1.08 (0.52, 2.26) | 0.84 | 1.12 (0.49, 2.56) | 0.79 |
| UPS (n=50) | 38.4 (21.3, 69.3) | 51.9 (31.2, 86.2) | 19.4 | NA | 0.79 | 0.98 (0.39, 2.44) | 0.96 | 1.28 (0.46, 3.60) | 0.64 |
| MFS (n=25) | 50.1 (31.5, 79.9) | 40.0 (13.7, 100.0) | NA | 13.4 | 0.53 | 1.49 (0.40, 5.63) | 0.55 | 3.29 (0.21, 51.63) | 0.40 |

Impact of ATRX loss on overall and disease-free survival among sarcoma patients in TCGA. Hazard ratios and corresponding p-values for overall survival and progression free survival obtained from cox-proportional hazard modeling. Model 1 was adjusted for age, sex, and histologic subtype (unless examined within a particular subtype). Model 2 was adjusted for age, sex, tumor size (> 10 cm, 5-10 cm, < 5 cm), and subtype (unless examined within a particular subtype). Median survival was not available for histologic subtypes, where overall or disease-free survival were greater than.

### Figures

**Figure S1.**
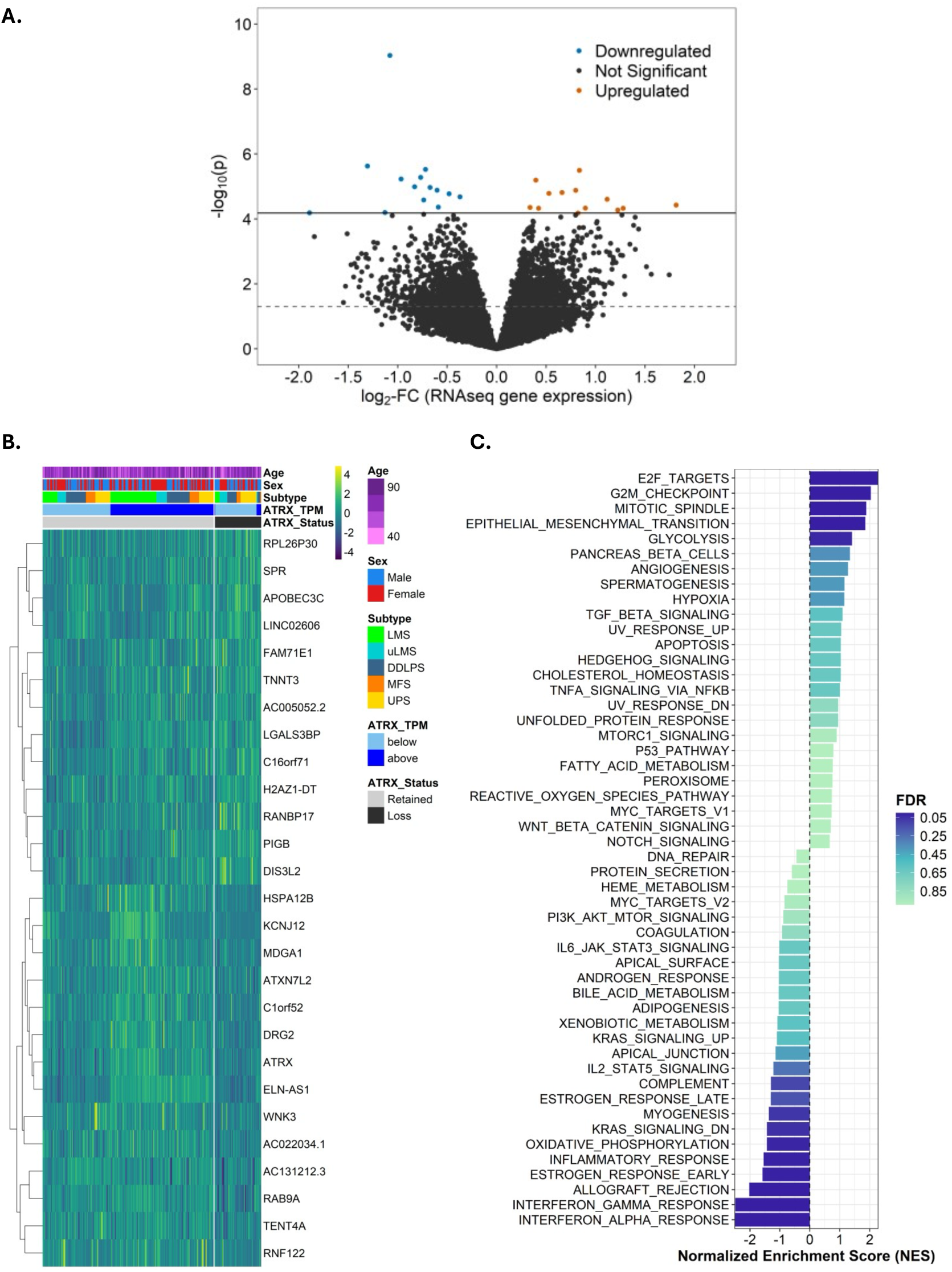
Impact of ATRX loss on global gene expression across all sarcomas. **A**) Volcano plot presents log_2_-fold change (log_2_-FC) estimates and –log_10_-transformed p-values across all gene expression in all subtypes. Estimates and p-values obtained from adjusted linear model examining the impact of ATRX loss on normalized gene counts. Models adjusted for histology site, age at diagnosis, and sex. Blue, red, and dark gray points correspond to gene expression changes that were downregulated in all sarcomas with ATRX loss (FDR < 0.05, log_2_-FC < 0), upregulated in all sarcomas with ATRX loss (FDR < 0.05, log_2_-FC > 0), and not associated with ATRX loss (FDR > 0.05), respectively. Solid and dotted line corresponds to FDR < 0.05 threshold and p < 0.05, respectively. **B**) Heatmap of gene expression associated with ATRX loss (FDR < 0.05, n=27 genes). Scaled, log_2_-transformed TPM values presented in plots and clusters identified using consensus clustering (see *Methods* for more details). **C**) Normalized enrichment scores (NES) across hallmark gene sets (see Methods). NES generated using GSEA approach and ranked gene list by log_2_-FC. NES plotted by size and direction and shaded by FDR of enrichment score p-value.

**Figure S2.**
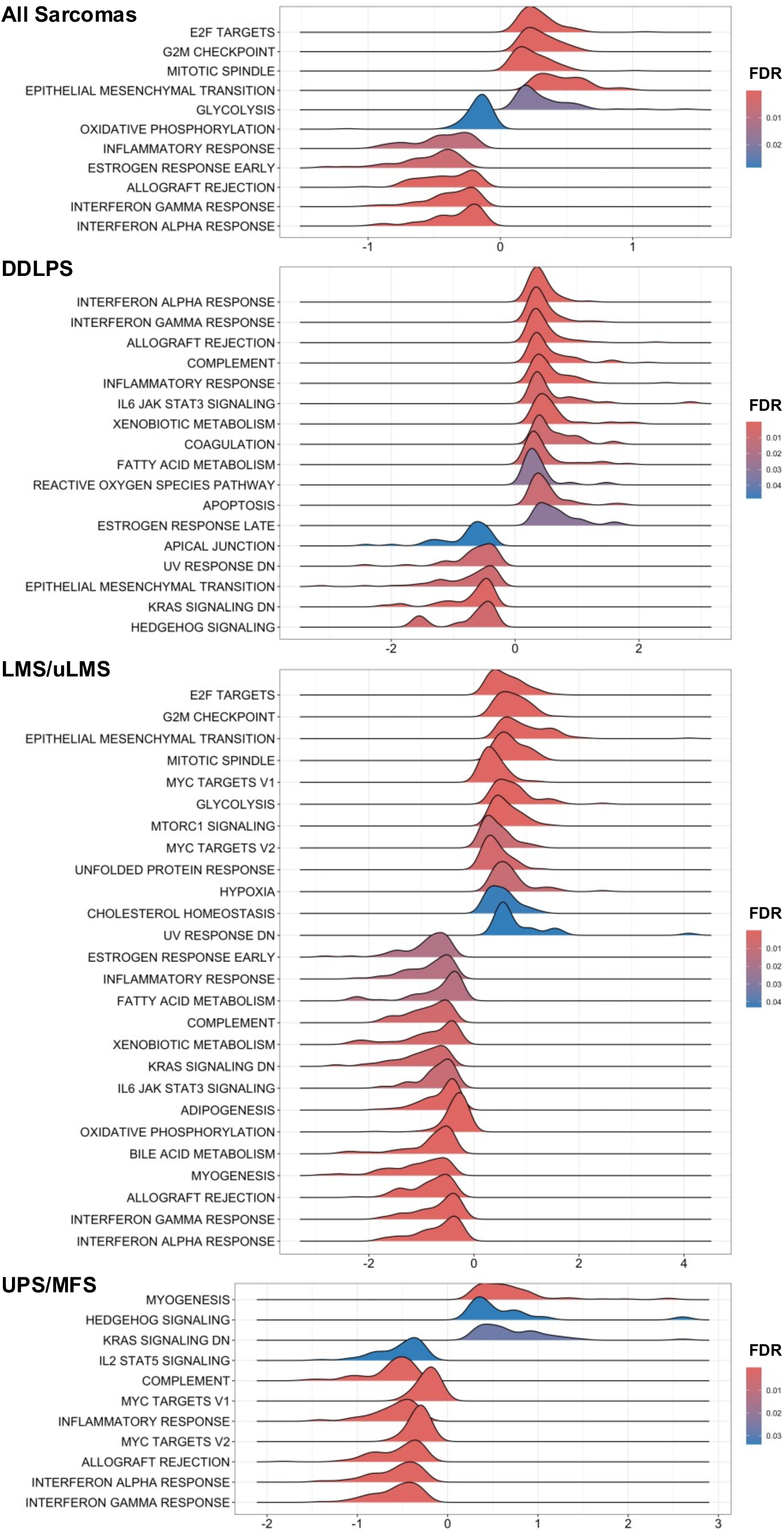
Distribution of log_2_-FC for genes within significantly enriched Hallmark gene sets. Ridgeplots show distribution of log_2_-FC for genes within Hallmark gene sets associated with ATRX loss (FDR < 0.05) across and within subtypes. X-axis shows value of log_2_-FC and fill color corresponds to adjusted p-value using Benjamini-Hochberg approach (FDR). Results from each histology subtype are provided.

**Figure S3.**
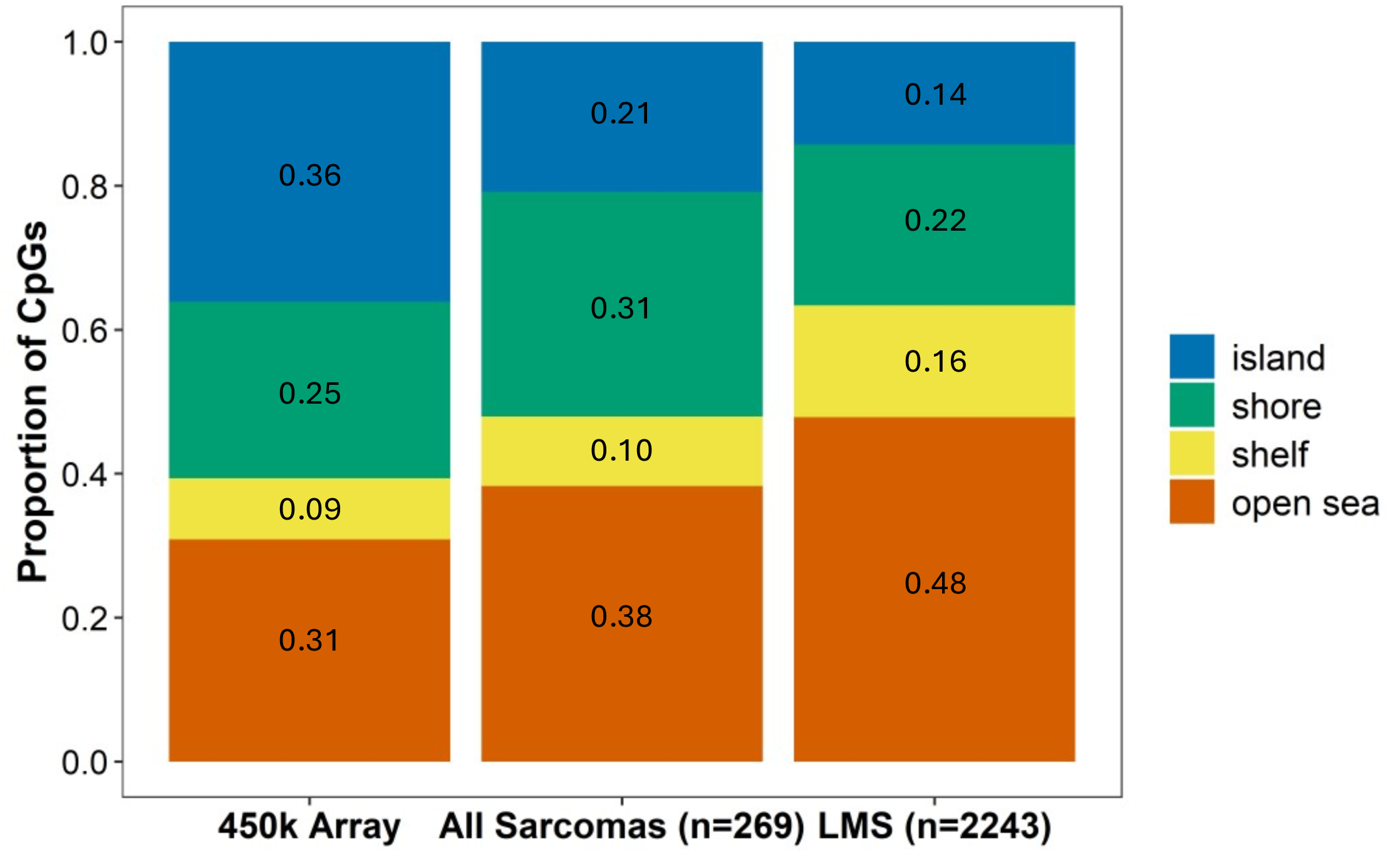
Distribution of differentially methylated CpGs associated with ATRX loss with relation to CpG island Proportion of CpGs annotating to island (high density of CpGs), shore (within 2 kilobases of island), shelf (within 5 kilobases from island), and open sea (lone CpGs).

**Figure S4.**
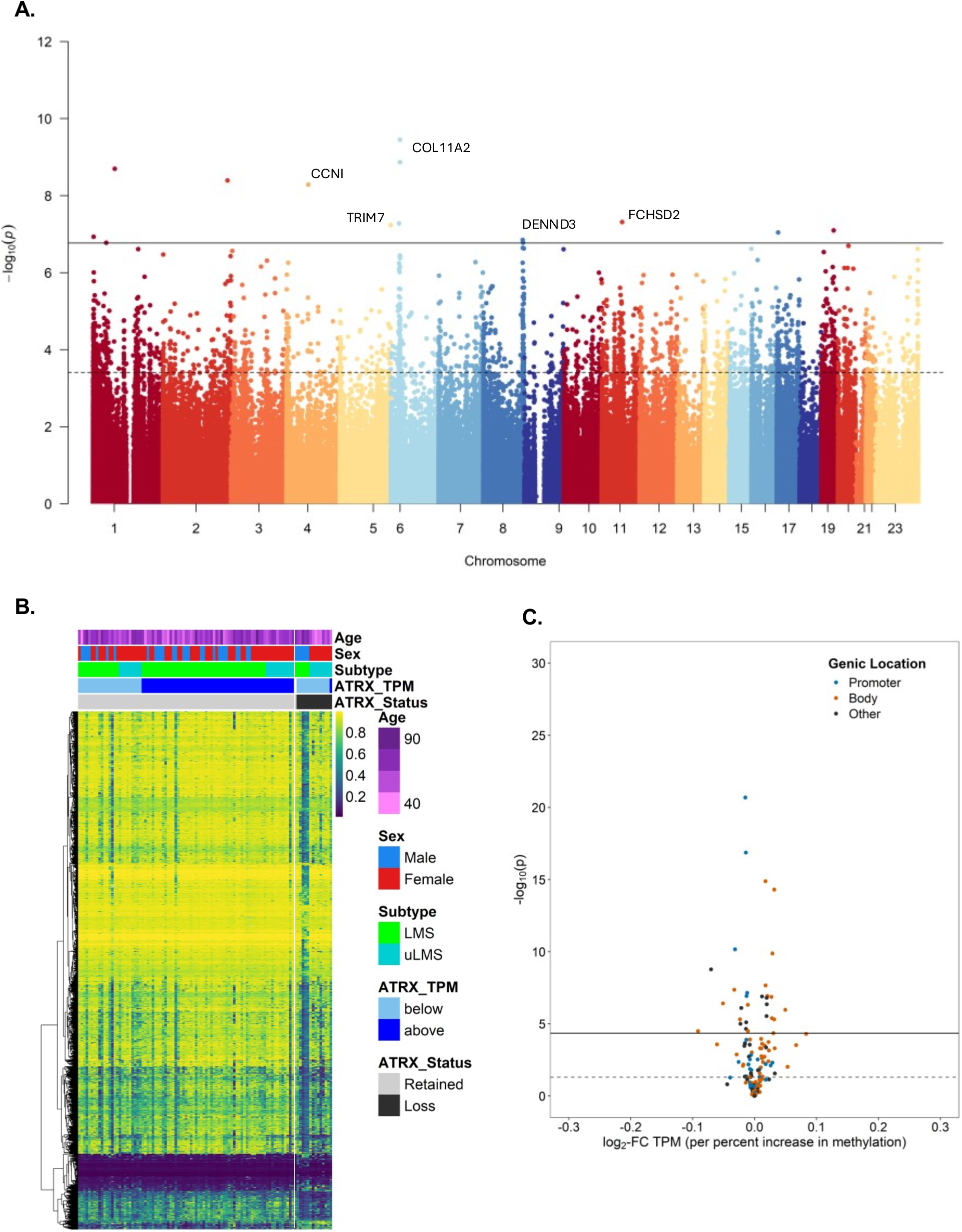
Impact of ATRX loss on genome-wide DNA methylation in LMS (all sites). A) Manhattan plot of associations between ATRX loss and CpG-specific methylation (M-values) from linear model adjusted for age, sex, and site (uterine vs. non-uterine) (see *Methods* for details). B) Heatmap of CpGs associated with ATRX loss in LMS (FDR < 0.05, n=2243). C) Association between ATRX-associated methylation and *cis*-gene expression.

**Figure S5.**
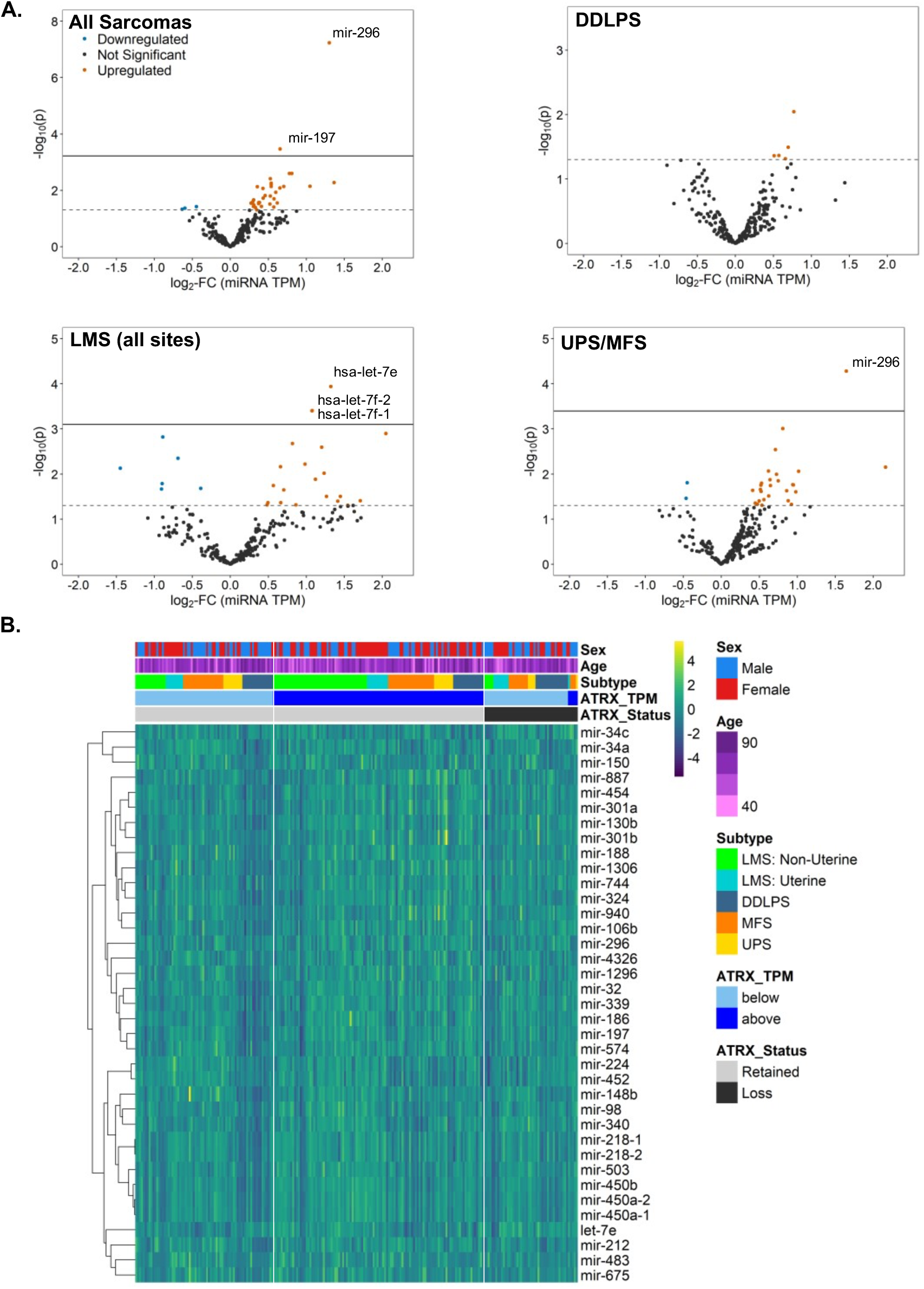
Impact of ATRX loss on miRNA expression in sarcomas. **A)** Volcano plot presents log_2_-fold change (log_2_-FC) estimates and –log_10_-transformed p-values across miRNA expressed in subtypes. Estimates and p-values obtained from adjusted linear model examining the impact of ATRX loss on log_2_-transformed TPM miRNA expression (see *Methods*) Blue, red, and dark gray points correspond to miRNA changes that were downregulated in LMS with ATRX loss (FDR < 0.05, log_2_-FC < 0), upregulated in LMS with ATRX loss (FDR < 0.05, log_2_-FC > 0), and not associated with ATRX loss (FDR > 0.05), respectively. Solid and dotted lines correspond to FDR < 0.05 threshold and p < 0.05, respectively. **B)** Heatmap of gene expression associated with ATRX loss (FDR < 0.05, n=94 genes). Scaled, log_2_-transformed TPM values presented in plots and clusters identified using consensus clustering (see *Methods* for more details).

**Figure S6.**
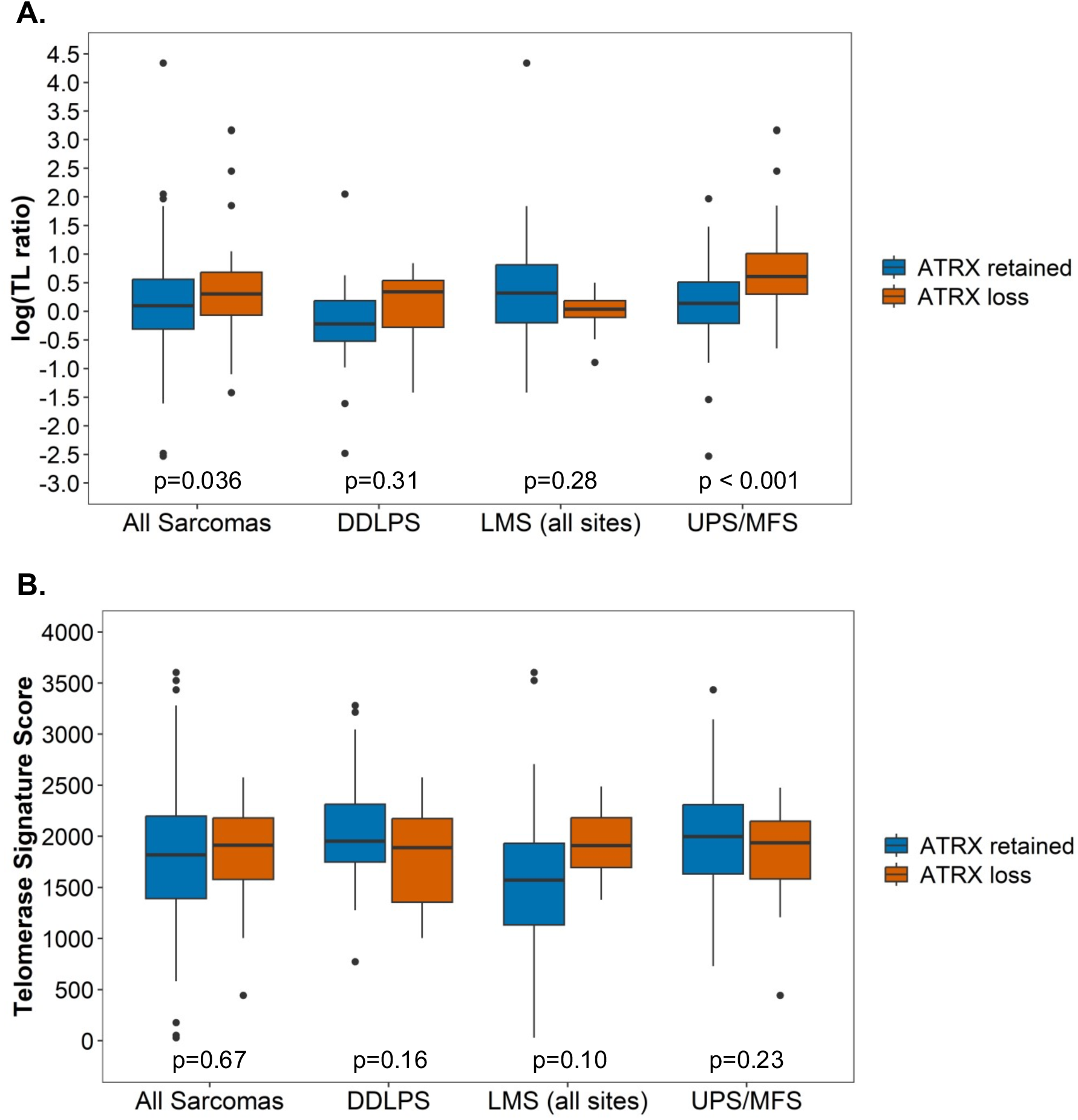
Relationship between telomere length, telomerase, and ATRX loss among sarcoma histologic subtypes. **A)** Telomere length ratio (TL ratio) is the tumor TL/normal tissue TL and was log-transformed for normality. TL was estimated from either whole exome sequencing (WXS) or whole genome sequencing (WGS) using TelSeq (see *Methods* and Barthel et al. 2017). P-values obtained from linear models adjusted for age, sex, histology (when relevant), and library type (WXS, WGS). For LMS, site (uterine, non-uterine) was also included in model. **B)** Telomerase signature score derived from RNA-seq gene expression (see *Methods* and Barthel et al. 2017). P-values obtained from linear models adjusted for age, sex, and histology (when relevant). For LMS, site (uterine, non-uterine) was also included in model.

**Figure S7.**
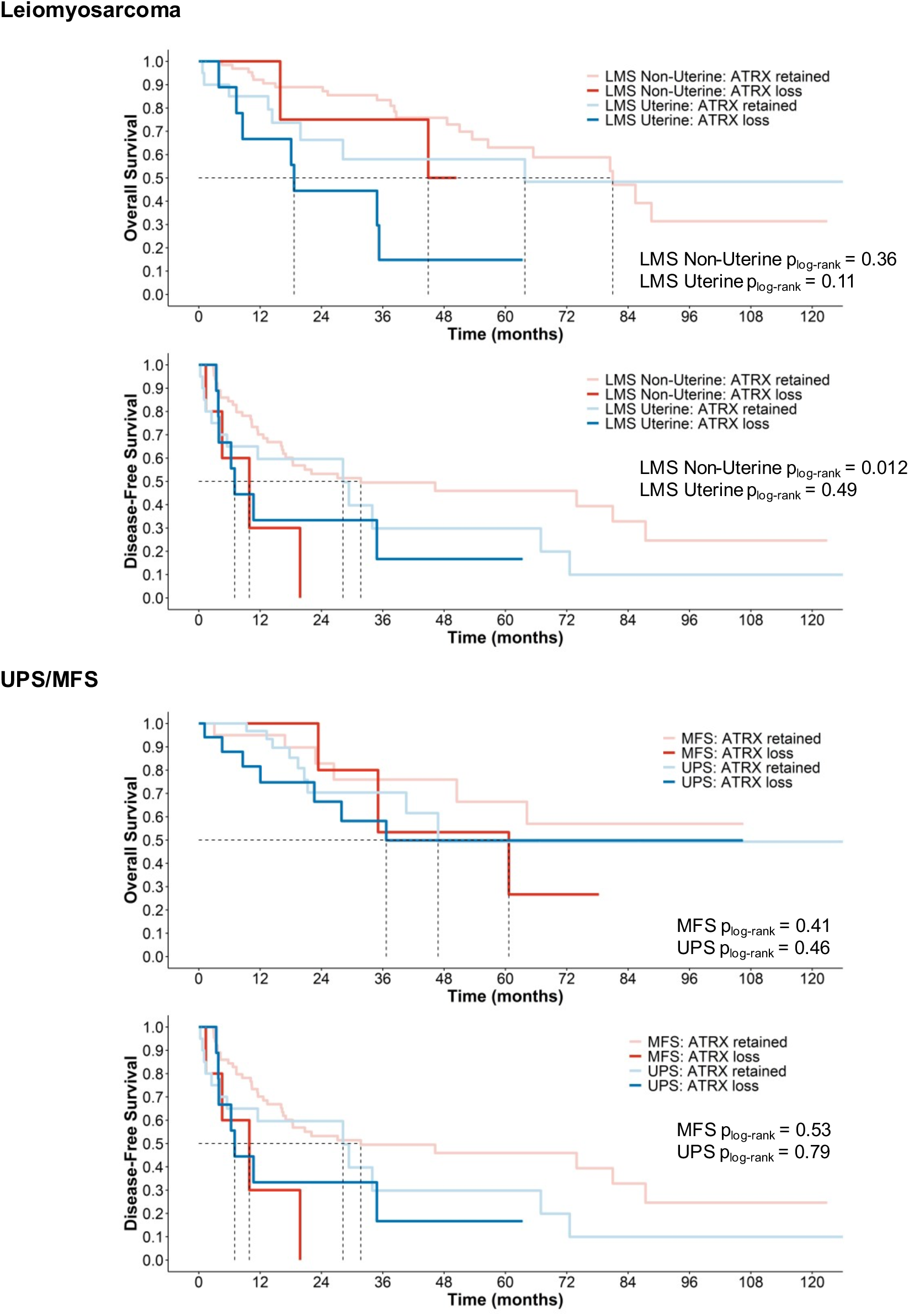
Overall and disease-free survival by subgroups of LMS, uLMS, UPS, and MFS. Kaplan-Meier plots of overall and disease-free survival stratified by ATRX status (loss and retained) and subgroup. Dashed lines correspond to median survival (in months). Log-rank tests were used to compare survival distribution by ATRX status within each subgroup and p-value is shown. Results from cox-proportional hazards modeling adjusted for age, sex, and tumor size, are shown in **Table 2**.

### Data Tables

**Data Table 1.** Associations between ATRX loss and gene expression (all genes).

**Data Table 2.** Over-representation analysis of differentially expressed genes associated with ATRX loss in LMS/uLMS.

**Data Table 3.** Associations between ATRX loss and methylation (p < 0.05).

**Data Table 4.** Differentially methylated regions associated with ATRX loss across all sarcomas.

**Data Table 5.** Associations between ATRX loss and methylation in LMS/uLMS (FDR < 0.05).

**Data Table 6.** Differentially methylated regions associated with ATRX loss across LMS (all sites)

**Data Table 7.** Impact of ATRX-associated methylation on cis-gene expression across all sarcomas.

**Data Table 8.** Impact of ATRX-associated methylation on cis-gene expression across LMS (all sites).

**Data Table 9.** Over-representation of genes in GO Biologic Process terms among genes whose expression is associated with differential methylation in sarcomas with ATRX loss.

**Data Table 10.** Over-representation of genes in GO Biologic Process terms among genes whose expression is associated with differential methylation in LMS/uLMS with ATRX loss.

**Data Table 11.** Associations between ATRX loss and transposable element expression.

**Data Table 12.** Associations between ATRX loss and miRNA expression.

